# Large Language Models forecast Patient Health Trajectories enabling Digital Twins

**DOI:** 10.1101/2024.07.05.24309957

**Authors:** Nikita Makarov, Maria Bordukova, Raul Rodriguez-Esteban, Fabian Schmich, Michael P. Menden

**Author notes:** Equal contribution (Nikita Makarov, Maria Bordukova).

## Abstract

**Background:** Generative artificial intelligence (AI) accelerates the development of digital twins, which enable virtual representations of real patients to explore, predict and simulate patient health trajectories, ultimately aiding treatment selection and clinical trial design. Recent advances in forecasting utilizing generative AI, in particular large language models (LLMs), highlights untapped potential to overcome real-world data (RWD) challenges such as missingness, noise and limited sample sizes, thus empowering the next generation of AI algorithms in healthcare.

**Methods:** We developed the Digital Twin - Generative Pretrained Transformer (DT-GPT) model, which utilizes biomedical LLMs using rich electronic health record (EHR) data. Our method eliminates the need for data imputation and normalization, enables forecasting of clinical variables, and preliminary explainability through a human-interpretable interface. We benchmarked DT-GPT on RWD including long-term US nationwide non-small cell lung cancer (NSCLC) and short-term Intensive Care Unit (ICU) datasets.

**Findings:** DT-GPT surpassed state-of-the-art machine learning methods in patient trajectory forecasting on mean absolute error (MAE) for both the long-term (3.4% MAE improvement) and the short-term (1.3% MAE improvement) dataset. Additionally, DT-GPT was capable of preserving cross-correlations of clinical variables (average R^2^ of 0.98), handling data missingness and noise. Finally, we discovered the ability of DT-GPT to provide insights into a forecast’s rationale and to perform zero-shot forecasting on variables not used during fine-tuning, outperforming even fully trained task-specific machine learning models on 13 clinical variables.

**Interpretation:** DT-GPT demonstrates that LLMs can serve as a robust medical forecasting platform, empowering digital twins which virtually replicate patient characteristics beyond their training data. We envision that LLM-based digital twins will enable a variety of use cases, including clinical trial simulations, treatment selection and adverse event mitigation.

## 1. Introduction

Clinical forecasting involves predicting patient-specific health outcomes and clinical events over time, which is of paramount importance for patient monitoring, treatment selection and drug development.^1^ Digital twins are virtual representations of patients that leverage a patient’s medical history to generate detailed multi-variable forecasts of future health states.^2^ The application of digital twins is poised to revolutionize healthcare in areas such as precision medicine, predictive analytics, virtual testing, continuous monitoring, and enhanced decision support.^3^

Generative artificial intelligence (AI) holds promise for creating digital twins due to its potential to produce synthetic yet realistic data, but this area of application is still in its infancy.^4^ Generative AI methods for predicting patient trajectories include recurrent neural networks, transformers and stable diffusion.^5–9^ These often fall short in terms of handling missing data, interpretability and performance. These challenges can be partially addressed by causal machine learning, but these algorithms face limitations related to small datasets or being confined to simulations.^10,11^

Recent breakthroughs in generative AI have been achieved with foundation models, which are pre-trained AI models adaptable to various specific tasks involving different types of data. Most foundation models for patient forecasting focus on single-point predictions rather than comprehensive longitudinal patient trajectories, which are needed for clinical decision-making.^12^ Less explored for this purpose remain text-focused Large Language Models (LLMs), which have demonstrated forecasting capabilities,^13,14^ including the ability of zero-shot forecasting, i.e. forecasting without any prior specific training in the task, thus highlighting their remarkable generalizability.^15,16^

We propose the creation of digital twins based on LLMs that leverage data from electronic health records (EHRs). EHRs are a key source of training data for machine learning models in healthcare, as they record patient characteristics such as demographics, diagnoses and lab results over time.^17^ However, they pose specific challenges such as data heterogeneity, rare events, sparsity and quality issues.^12^ There have been developments in machine learning to overcome these challenges, especially for data sparsity, usually by adapting the model’s architecture, resulting in increased model complexity and the introduction of further assumptions on the data.^6,9^

We hypothesize that LLMs will empower digital twins and overcome these challenges. Here, we introduce the Digital Twin - Generative Pretrained Transformer (DT-GPT) model (**Fig. 1**), which enables: i) forecasting of clinical variable trajectories, ii) zero-shot predictions of clinical variables not previously trained on, and iii) preliminary interpretability utilizing chatbot functionalities. We analyze the performance of the model by forecasting laboratory values on both a long-term scale (up to 13 weeks) for non-small cell lung cancer (NSCLC) patients, as well as a short-term scale (next 24 hours) for Intensive Care Unit (ICU) patients. Based on our results, we anticipate that DT-GPT will pave the way for AI-based digital twins in healthcare.

**Figure 1:**
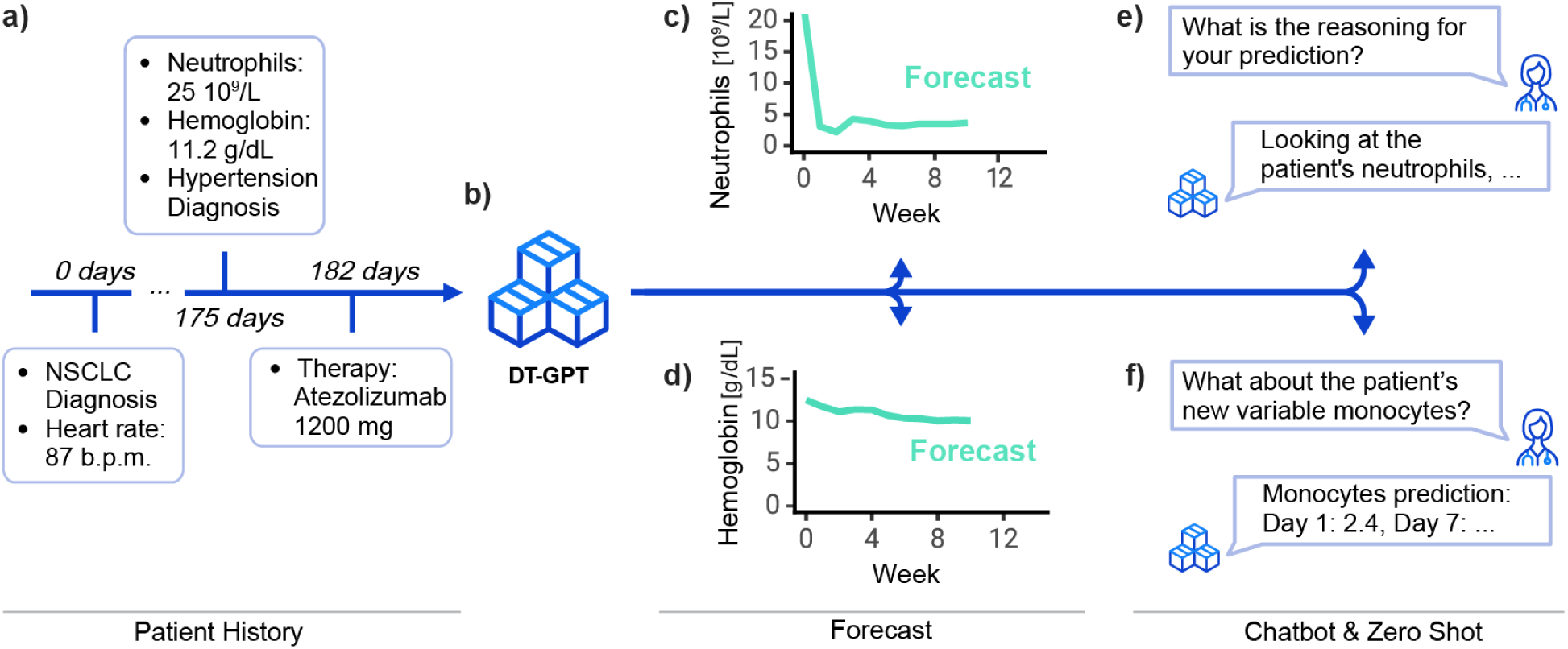
The LLM-based DT-GPT framework enables forecasting patient trajectories, identifying key variables, and zero-shot predictions. Here exemplified, **a**) a sparse synthetic patient timeline, which **b**) DT-GPT utilizes for generating longitudinal clinical variable forecasts, e.g., **c**) neutrophil and **d**) hemoglobin blood levels. DT-GPT can **e**) chat and respond to inquiries about important variables, as well as **f**) perform zero-shot forecasting on clinical variables previously not used during training.

## 2. Methods

DT-GPT is a method that employs pre-trained LLMs fine-tuned on clinical data (**Fig. 2a)**. Notably, DT-GPT is agnostic regarding the underlying LLM and can be applied without architectural changes to any general-purpose or specialized text-focused LLM.

**Figure 2:**
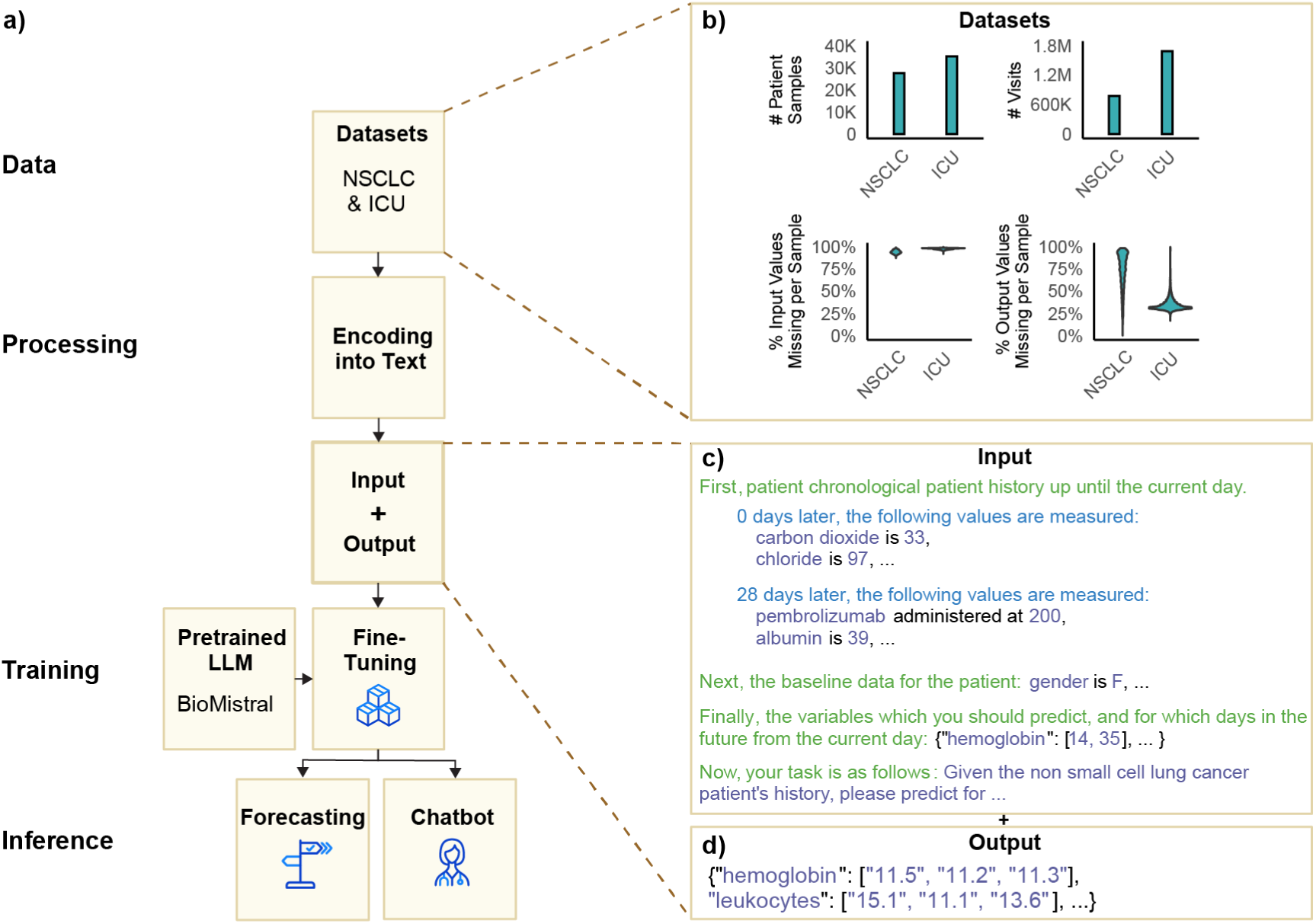
The DT-GPT framework transforms EHRs into text and subsequently fine-tunes an LLM on this data. **a**) Overview of the pipeline: datasets are split and encoded into input/output text based on landmark timepoints, then used to fine-tune an LLM, here BioMistral. The model output is evaluated for trajectory forecasting whilst zero-shot predictions and variable importances are explored via a chat interface. **b)** Sample size, visit frequency, and sparsity of the non-small cell lung cancer (NSCLC) and intensive care unit (ICU) datasets. **c)** Input and **d)** output encoded examples, emphasizing the chronological encoding of observations.

### 2.1 Datasets and data preparation

We trained and evaluated DT-GPT for forecasting patients’ laboratory values across two independent datasets, namely long-term and short-term trajectories of non-small cell lung cancer (NSCLC) and intensive care unit (ICU) patients, respectively. For the US-based NSCLC dataset, we used the nationwide Flatiron Health EHR-derived de-identified database. The data are de-identified and subject to obligations to prevent re-identification and protect patient confidentiality. The Flatiron Health database is a longitudinal database, comprising de-identified patient-level structured and unstructured data, curated via technology-enabled abstraction.^18,19^ During the study period, the de-identified data originated from approximately 280 cancer clinics (∼800 sites of care).

The study included 16,496 patients diagnosed with NSCLC from 01 January 1991 to 06 July 2023. The majority of patients in the database originate from community oncology settings; relative community/academic proportions may vary depending on study cohort. Patients with a birth year of 1938 or earlier may have an adjusted birth year in Flatiron Health datasets due to patient de-identification requirements. To harmonize the data, we aggregated all values in a week based on the last observed value. We focused on the 50 most common diagnoses and 80 most common laboratory measurements, complemented by the Eastern Cooperative Oncology Group (ECOG) score, metastases, vitals, drug administrations, response and mortality variables totaling 773,607 patient-days across 320 variables.

##### Research in Context

###### Evidence before this study

Digital Twins (DTs) for forecasting patient-specific clinical trajectories, events and outcomes are being increasingly realized by the means of generative artificial intelligence (AI). We conducted a comprehensive search using Google Scholar and Scopus for studies and reviews on methods for predicting longitudinal patient trajectories, published in English between Jan 1, 2019, and March 31, 2024. The search employed the following keyword combinations: “forecasting” and “patient trajectory”, “forecasting” and “patient”, “forecasting” and “clinical”, “prediction” and “patient trajectory” and “time”, “forecasting and electronic health records”, “prediction” and “patient” and “time”, “prediction” and “clinical” and “time”, “prediction” and “patient trajectory” and “longitudinal”, “prediction” and “patient” and “longitudinal”, “prediction” and “clinical” and “longitudinal”, “foundation model” and “electronic health records”, “large language model” and “electronic health records”. We restricted the search results to the studies describing generative AI-based methods. The identified studies introduce high-performing clinical forecasting methods but their validation is limited to a single application only with a specific disease indication or dataset. Additionally, these methods typically require extensive data preprocessing, such as imputation of missing values and normalization. While large language models and foundation models offer a more general setup applicable to various research questions and datasets, existing methods predominantly focus on single time-point predictions rather than longitudinal predictions. Moreover, the interpretability of existing models is limited.

###### Added value of this study

This study introduces the Digital Twin - Generative Pretrained Transformer (DT-GPT) model, a novel method to fine-tune large language models (LLMs) to forecast multi-variable patient trajectories, combining the advantages of existing methods while overcoming the limitations of data heterogeneity and sparsity associated with electronic health records (EHRs). DT-GPT achieves state-of-the-art forecasting performance on long-term US nationwide non-small cell lung cancer datasets and short-term intensive care unit datasets, demonstrating its applicability to multiple disease conditions with various time horizons and both regular and irregular time sampling. Furthermore, DT-GPT learns relationships and preserves cross-correlation between variables, enabling zero-shot (i.e., without any training) prediction of clinical variables previously not trained on. Finally, the interactive interface provides preliminary prediction explainability through chatbot functionality.

###### Implications of all the available evidence

Generative AI-based models enhance the capabilities of patient DTs for treatment selection, patient monitoring, and clinical trial support by creating state-of-the-art patient trajectory predictions. DT-GPT advances the development of DTs by reducing the need for extensive data preprocessing and enabling interaction with the model through a human-interpretable interface. Our method is anticipated to be easily accessible to clinicians, allowing efficient simulations of patient trajectories under various scenarios to support clinical decision-making.

For every NSCLC patient, we divided their trajectory into input and output segments based on the start date of each line of therapy to create each patient sample. All variables up to the start date were considered input data. The objective was to predict the weekly values up to 13 weeks after the start date of the following variables and their respective LOINC codes: hemoglobin (718-7), leukocytes (26464-8), lymphocytes/leukocytes (26478-8), lymphocytes (26474-7), neutrophils (26499-4) and lactate dehydrogenase (2532-0). These variables were selected due to their frequent measurement and relevance in reflecting key characteristics of NSCLC treatment response **(Appendix A1)**.

To demonstrate the generalizability of DT-GPT, we analyzed ICU trajectories from the publicly-accessible Medical Information Mart for Intensive Care IV (MIMIC-IV) dataset.^20^ We employed an established processing pipeline, resulting in 300 input variables across 1,686,288 time points from 35,131 patients.^21^ Here, the objective was to predict a patient’s future hourly lab variables given their first 24 hours in the ICU. Specifically, the patient history was considered as the first 24 hours for all variables, and the task was to forecast the future 24 hourly values for the following variables: O2 saturation pulse oximetry, respiratory rate and magnesium. These variables were selected due to their clinical relevance, high temporal variability, and the fact that at least 50% of patients had at least one measurement for each.

These criteria not only increased the forecasting challenge, but also ensured wide representation across the patient population.

Both datasets were randomly split at the patient level into 80% training, 10% validation, and 10% test set. Thus, each set comprised disjoint sets of patients to avoid data leakage. The test sets were solely used for final evaluation and to assess the model’s generalizability (**Fig. 2b**; **Appendix A1)**.

### 2.2 Encoding

We encoded patient trajectories by using templates that converted medical histories based on EHRs into a text format compatible with LLMs, as proposed by *Xue et al.*^14^ and *Liu et al*.^15^ (**Fig. 2c,d**; **Appendix A4)**. The input template is structured into four components: 1) Patient history, 2) demographic data, 3) forecast dates and 4) prompt. The patient history contains a chronological description of patient visits, requiring no data imputation for missing variables. The output trajectories were also encoded using templates, containing only the relevant output variables for the forecasted time points. We utilized a manually developed template for input encoding and JSON-format encoding for the output (**Appendix A4, A10)**.

### 2.3 LLMs and fine-tuning

We utilized the biomedical LLM BioMistral 7B DARE, since it is provided with an open source license and based on a recognized LLM.^22^ Furthermore, BioMistral is instruction tuned and through its biomedical specialization incorporates compressed representations of vast amounts of biomedical knowledge. We further fine tuned this LLM using the standard cross entropy loss, masked so that the gradient was only computed on the output text. We performed 30 predictions for each patient sample during evaluation, then took the mean for each time point as the final prediction.^16,23^ All hyperparameters used in fine-tuning are shown in **Appendix A5**.

### 2.4 Chatbot and zero-shot learning

We employed the DT-GPT model to run a chatbot based on patient histories for prediction explanation and zero-shot forecasting. For this, first we used DT-GPT to generate forecasting results from patient history and, consecutively, added a task-specific prompt surrounded by the respective instruction-indication tokens to the DT-GPT chat history for receiving a response. For prediction explanation, the prompt asked for the most important variables influencing the predicted trajectory. For zero-shot forecasting, the prompt specified the output format and days to predict new clinical variables that were not subject to optimization during training. Example prompts and chatbot interactions for both tasks are provided in **Appendix A6** and **Fig. 5a,e**.

### 2.5 Baseline models

We employed five multi-step, multivariate baselines, ranging from a simple baseline to state-of-the-art forecasting models. Specifically, we used a naïve model that copies over the last observed value, a linear regression model, a time series LightGBM model, a Temporal Fusion Transformer as well as a TiDE model.^8,24,25^ These models were selected due to their ability to handle future variables and state-of-the-art performance in both medical and standard time series forecasting.^26,27^ The hyperparameters and training details are shown in **Appendix A7**.

### 2.6 Evaluation

For evaluation of the forecasted patient trajectories, we used the mean absolute error (MAE) as our primary metric. We first standardized and calculated the pairwise error between the forecasted value and the true value of a given sample and time step, averaged over all patients and timepoints, and then averaged again over all variables.^25^ All error bars refer to the standard error of the model aggregated across all variables.^28^ For the evaluation of the effects of RWD missingness we randomly sampled 200 patients of the test set. Chatbot exploration and zero-shot forecasting were analyzed on the whole test set.

## 3. Results

DT-GPT achieved state-of-the-art forecasting performance, being stable to address common RWD challenges and forecast zero-shot lab variables. Additionally, we explored how DT-GPT provides preliminary insights into its predictions.

### 3.1 DT-GPT achieved state-of-the-art forecasting performance

DT-GPT achieved the lowest overall mean absolute error (MAE) across both benchmark tasks (**Table 1**). On the NSCLC dataset, DT-GPT achieved an average MAE of 0·55 ± 0·04, whilst LightGBM, the second best model, achieved an average MAE of 0·57 ± 0·05, showing a relative improvement of 3·4% (**Table 1a**). On the ICU dataset, DT-GPT achieved an average MAE of 0·59 ± 0·03, whilst the second best model, LightGBM, performed at 0·60 ± 0·03, equivalent to a 1.3% improvement (**Table 1b; Appendix A8**).

**Table 1:** Benchmark of clinical variable forecasting across two datasets. DT-GPT outperformed the baselines in the majority of cases on both **a**) the long-term non-small cell lung cancer (NSCLC) and **b**) the short-term intensive-care unit (ICU) datasets. All errors refer to mean absolute error (MAE; lower is better) normalized by standard deviation. For example, an MAE of 0·6 means that the model prediction is on average within 0·6 standard deviations of the observed value. Best performing models are highlighted in bold.

| Model | a) NSCLC |  |  |  |  |  | b) Intensive Care Unit |  |  |
| --- | --- | --- | --- | --- | --- | --- | --- | --- | --- |
|  | Hemo-<br>globin | Leuko-<br>cytes | Lympho-<br>cytes/<br>Leuko-<br>cytes | Lympho-<br>cytes | Neutro-<br>phils | Lactate<br>Dehydro-<br>genase | Magne-<br>sium | Resp.<br>Rate | Oxygen<br>Saturation |
| Copy Forward | 0.698 | 0.969 | 0.731 | 0.569 | 0.974 | 0.433 | 0.681 | 0.769 | 0.746 |
| Linear Regression | 0.486 | 0.782 | 0.668 | 0.506 | 0.778 | 0.475 | 0.606 | 0.680 | 0.681 |
| Temporal Fusion Transformer | 0.469 | 0.719 | 0.651 | 0.463 | 0.717 | 0.480 | 0.537 | 0.635 | 0.644 |
| TiDE | 0.464 | 0.737 | 0.655 | 0.465 | 0.740 | 0.453 | 0.534 | 0.635 | 0.652 |
| LightGBM | 0.453 | 0.727 | **0.644** | 0.456 | 0.734 | 0.425 | 0.520 | **0.634** | 0.644 |
| DT-GPT (ours) | **0.440** | **0.689** | 0.650 | **0.437** | **0.699** | **0.417** | **0.505** | 0.636 | **0.635** |

DT-GPT forecasts preserved inter-variable relationships. The correlations between the variables forecasted by DT-GPT aligned with the correlations between the variables in the test datasets with an R^2^ of 0·98 and 0·99, whilst those of LightGBM achieved an R^2^ of 0·97 and 0·99 (**Appendix A8**) on the NSCLC and ICU datasets, respectively. Additionally, DT-GPT outperformed LightGBM in the majority of timepoints in both datasets, demonstrating that the improvement was consistent across time (**Fig. 3a,b; Appendix A8**).

**Figure 3:**
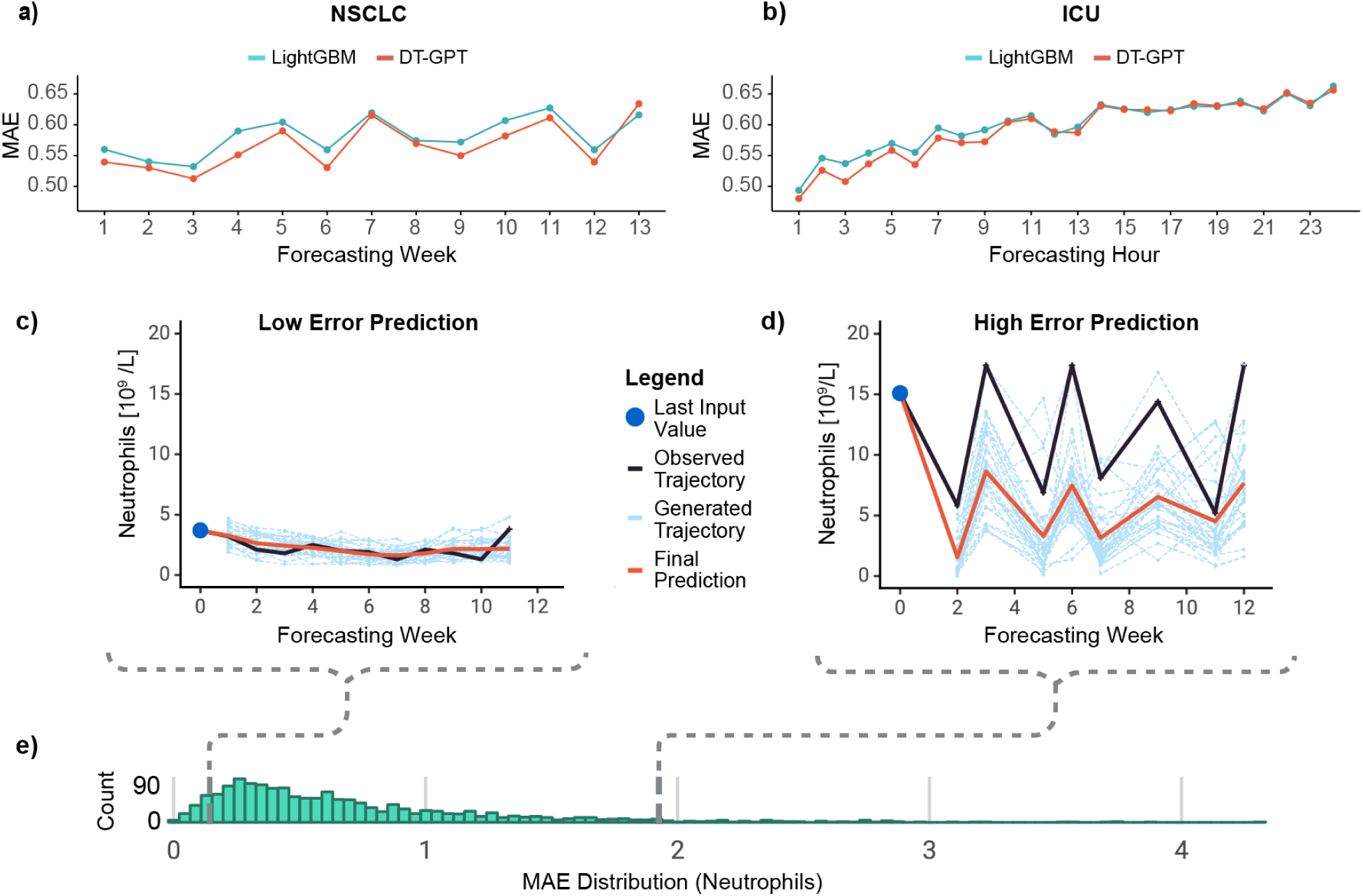
Performance analysis of DT-GPT. **a**) The long-term non-small cell lung cancer (NSCLC) and **b**) the short-term intensive-care unit (ICU) dataset, with the x-axis showing relative time points and the y-axis the corresponding mean absolute error (MAE), comparing with the second best forecasting model LightGBM. Here exemplified, DT-GPT forecasts of neutrophil counts in patients with **c**) low and **d**) high error, for all weeks where the ground truth exists. **e)** Histogram of MAE distribution for all predicted neutrophil counts.

DT-GPT can be further improved by exploring alternative trajectory aggregation methods. To inspect both low and high MAE predictions from DT-GPT, we visualized two sample individual-patient forecasts for the variable neutrophils (**Fig. 3c,d**) picked from the low and high end of performance distribution (**Fig. 3e**). Note that the final prediction was derived by averaging 30 generated trajectories and that, even in poor performing cases, individual non-averaged forecasted trajectories were sometimes able to capture parts of the true trajectory. To explore the importance of trajectory aggregation, we calculated the error given an optimal aggregation. To this end, we selected the individual trajectories with the lowest MAE and recalculated the hypothetical MAE on the NSCLC dataset, achieving a 26% improvement in error to 0·40 ± 0.02, without any further model training. Note that this is a theoretical lower bound. Finally, we noted that in the distribution of MAE for neutrophils across all patients, most of the errors were right-skewed, indicating that high errors came from a small number of outlier patients (**Fig. 3e**).

### 3.2 DT-GPT is robust to common RWD challenges

DT-GPT is flexible and robust to common practical data challenges, exhibiting desired properties in a variety of ablation studies, here exemplified on the average performance on all six clinical variables of the NSCLC dataset. First, DT-GPT performance was competitive with baselines after training with data corresponding to 5,000 patients and it further improved with the number of patients in the training dataset (**Fig. 4a**; **Table 1**). Additionally, DT-GPT could handle increased input missingness, with performance degradation only showing after more than 20% of the input was randomly masked, on top of the 94·4% initial missingness of the NSCLC dataset (**Fig. 4b**). Thirdly, DT-GPT was stable to misspellings in the input, only significantly degrading in performance after 25 misspellings per patient sample (**Fig. 4c**). Note that misspellings cannot be handled by most established machine learning methods and either require completely dropping or manual curation of the data.

**Figure 4:**
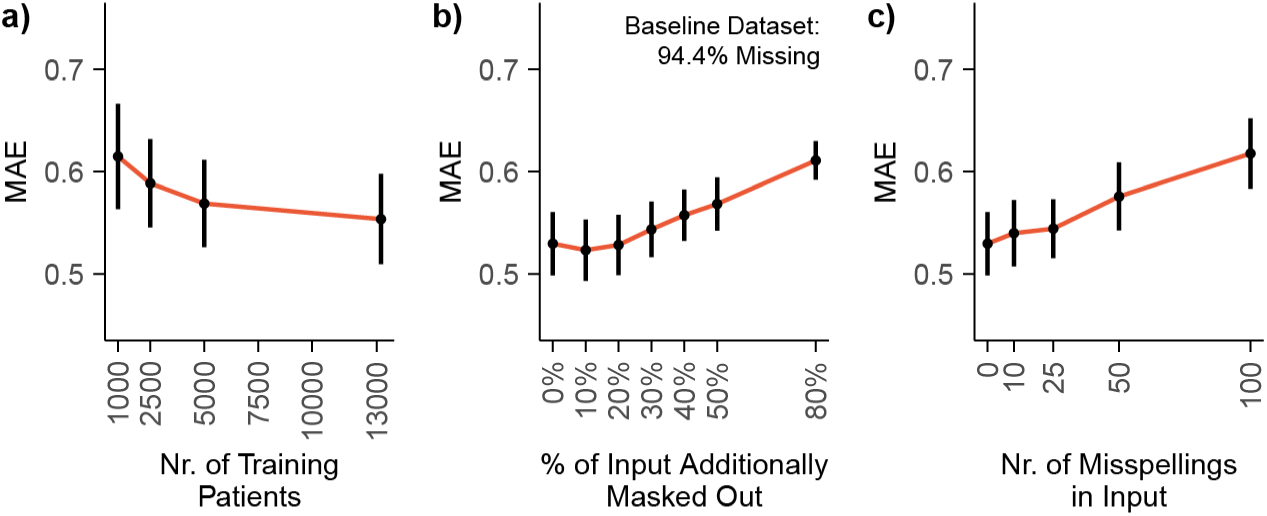
DT-GPT is robust to common RWD issues in the long-term NSCLC dataset. **a)** Mean absolute error (MAE) according to the number of patients in the training set. Assessing impact on MAE based on **b)** added missingness, on top of the baseline 94·4% missingness of the NSCLC dataset, and **c)** injected misspellings in the input.

### 3.3 DT-GPT enables prediction insights and zero-shot forecasting

DT-GPT retained its conversational capability post-fine-tuning for the forecasting task, facilitating user interaction and enabling the inquiry of prediction reasoning. For each patient sample, 10 predicted trajectories were generated, accompanied by a set of explanatory variables elucidating these predictions (**Fig. 5a**). We extracted explanatory variables from 25,575 out of 27,730 chatbot responses. The most influential variables were therapy, ECOG status and leukocyte count (**Fig. 5b; Appendix A11**). For example, the therapy significantly influenced hemoglobin dynamics: patients receiving immunotherapy and targeted therapy (EGFR inhibitors) generally exhibited higher hemoglobin levels over time compared to those undergoing chemotherapy or combination therapies (i.e. chemotherapy and immunotherapy), whose hemoglobin levels tended to decline due to the chemotherapy impacting bone marrow (**Fig. 5c; Appendix A12,A13**).^29^ Additionally, the last recorded ECOG value in a patient’s medical history impacted the predicted hemoglobin trajectories (**Fig. 5e**), with lower ECOG values (less performance restriction) correlating with higher hemoglobin levels over time, which is concordant with literature as well.^30^ These findings are also consistent with the original data (**Fig. 5d,f**).

**Figure 5:**
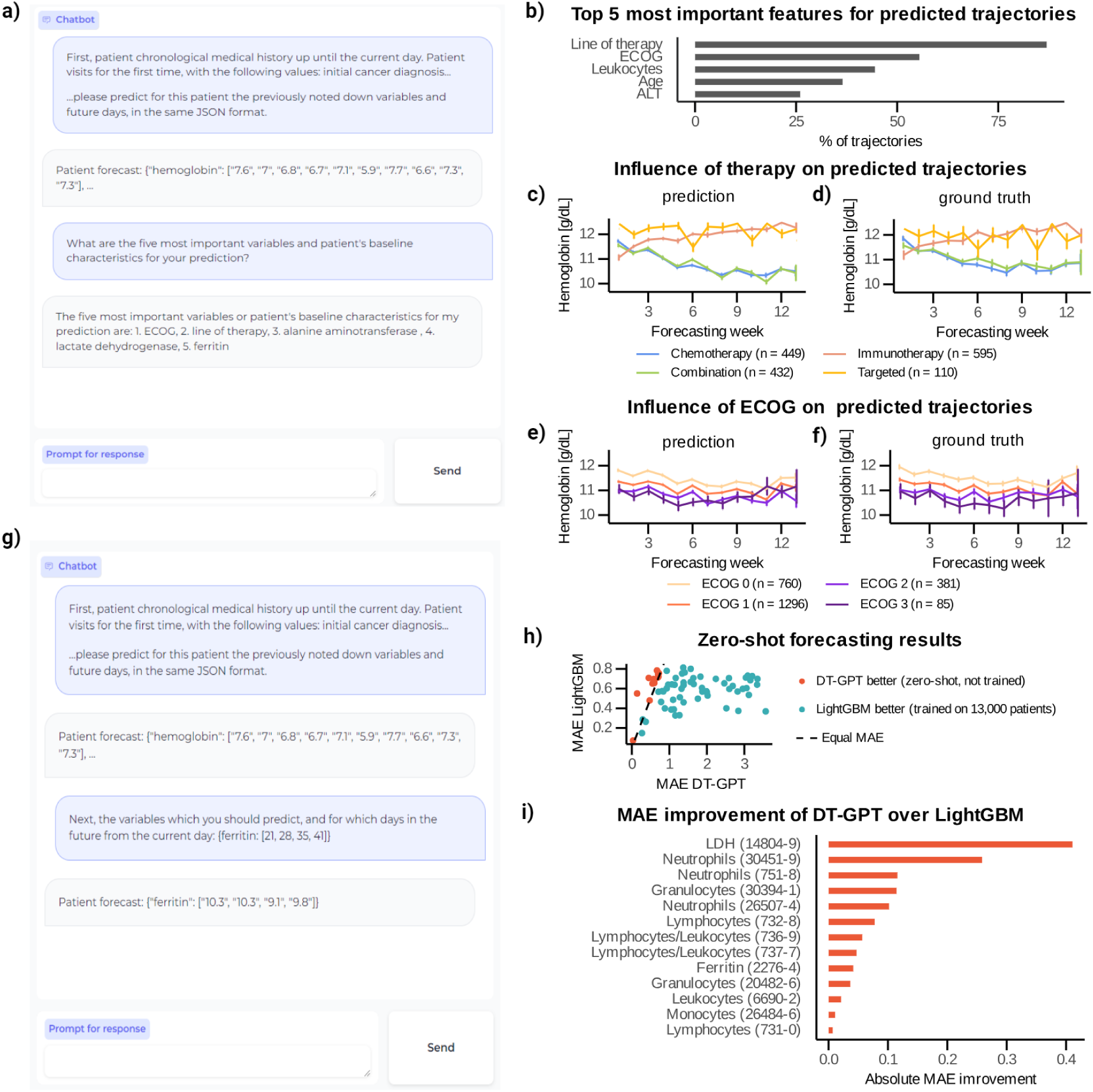
DT-GPT preserves its conversational ability after the fine-tuning, allowing inquiring into prediction rationale and zero-shot forecasting. **a)** Example of a chatbot interaction providing explanations for predictions. **b)** Five most important variables for predicting all variables derived from forecasting test patient samples with 10 predicted trajectories each. **c)** The most important variable, therapy, influences predicted hemoglobin trajectories, with **d)** the corresponding ground truth. Here, the lines show average trajectories and the error bars correspond to the standard error. **e)** The second most important variable, ECOG, influences predicted hemoglobin trajectories, and **f)** showing the corresponding ground truth. Lines represent average trajectories and the error bars correspond to the standard error. **g)** Example of a chatbot interaction for forecasting a variable not previously trained on. **h)** DT-GPT outperforms LightGBM models on 13 out of 69 non-target variables. Notably, LightGBM models were trained on more than 13,000 patient data in a supervised manner, whereas DT-GPT is performing here fully zero-shot predictions. **i)** DT-GPT is superior for variables more closely related to the target variables used during fine tuning, with the respective LOINC codes depicted in parentheses. (Abbreviations: ECOG - Eastern Cooperative Oncology Group performance status scale, LDH - Lactate dehydrogenase, ALT - Alanine aminotransferase).

DT-GPT supports zero-shot forecasting of 69 non-target clinical variables that are observed in patient medical histories but were not subject to model fine tuning. In our experiments, we forecasted each non-target variable separately (**Fig. 5g**) and extracted 81,004 trajectories from 81,918 forecasting results.

We compared the performance of zero-shot DT-GPT with a supervised LightGBM model, which was trained on each non-target variable separately using data from over 13,000 patients. Notably, LightGBM is a fully supervised model and therefore anticipated to perform better than a zero-shot model.

Surprisingly, however, zero-shot DT-GPT outperformed LightGBM on 13 out of 69 non-target variables (**Fig. 5h,i**). The variables with improved performance can be described as closely related to the target variables (**Fig. 5i**). For instance, *segmented neutrophils*, *band form neutrophils* and *neutrophils by automated count* have different LOINC codes from the trained variable (30451-9, 26507-4, 751-8, respectively), but are correlated with the target variable *neutrophils* (LOINC 26499-4). A table containing MAE values for DT-GPT and the LightGBM baseline is provided in **Appendix A14**.

## 4. Discussion

Our main finding is that a simple yet effective method allows training LLMs on EHRs to generate detailed patient trajectories that preserve inter-variable correlations. This method achieves novel zero-shot performance, potentially making DT-GPT a digital twin platform that can mimic individual patients, with applications such as treatment selection and clinical trial support.

Building on past LLM research in general forecasting, DT-GPT outperforms existing baselines in NSCLC and ICU datasets.^15,16^ These findings align with recent LLM forecasting developments, demonstrating that clinically-specific adjustments enable accurate predictions.^13,14^ Additionally, DT-GPT’s generative nature allows for multiple trajectory simulations per patient, offering insights into possible patient scenarios, cohort simulations and uncertainty estimates.

The positive performance of LLMs for patient forecasting may stem from parallels between natural language and biomedical data, such as non-random missingness. For example, a doctor might skip measuring blood pressure if a patient appears healthy, indicating information by omission. Natural language implicitly handles such ambiguity; unspoken words can still convey meaning or none at all. Recent advancements suggest that LLMs can capture these complex relationships.^31^

DT-GPT addresses electronic health record (EHR) challenges including noise, sparsity and lack of data normalization.^12^ Unlike most established machine learning models that require data normalization and imputation, DT-GPT operates without these requirements. Here, we demonstrated its robustness to sparsity, misspellings and noisy medical data often encountered in real-world datasets. Moreover, EHR data often contain mixed data encodings; for instance, drug information may vary in encoding, such as the dosage used or noted only as “administered”, both of which DT-GPT handles without additional preprocessing. Overall, DT-GPT simplifies and streamlines data preparation, thus enabling faster deployment across diverse datasets.

DT-GPT can be inquired about the rationale of predictions, which increases the interpretability of the model. This capability bridges the gap between medical expert and model, enabling the exploration of prediction rationales and alternative patient scenarios efficiently. We anticipate that this advancement represents a significant leap in human-computer interaction with AI predictions, which is expected to profoundly influence clinical practices in the near future.

DT-GPT enables zero-shot predictions, demonstrating its ability to forecast variables not explicitly included in its fine-tuning phase by learning their dynamics and adapting to novel tasks. Remarkably, zero-shot DT-GPT outperforms a supervised, fully-trained machine learning model on a subset of clinical variables, highlighting the pioneering potential of LLM-based approaches in RWD forecasting. This underscores the transformative potential of LLM-based models like DT-GPT to revolutionize forecasting in healthcare, suggesting a promising future for AI-driven advancements in clinical decision-making and precision medicine.

A challenge of LLM-based models is the restricted number of simultaneously forecasted variables. The current constraint on the number of forecasted variables is due to the limited sequence length of both input and output of the LLMs used in fine-tuning. Advances in extending the context length will enable modeling of additional patient variables. Furthermore, we anticipate that transitioning from zero-shot to few-shot learning, where the model receives further training on a small subset of data, would enable a wider span of forecasted variables and extend DT-GPT’s applicability to broader clinical challenges.

Another established shortcoming of LLM-based models is their tendency to hallucinate. In our case, this could be reflected in explainability results not necessarily providing true answers. This is critical for the medical domain and an active field of research in explainable AI, which we believe will be the focus of the next generation of LLM-based models.

Finally, we observe that high error predictions often occur due to the high variance between the multiple generated trajectories of each patient sample, with the mean aggregation into the final prediction not capturing key dynamics. It is thus an open challenge to develop improved aggregation methods, for example by using a second LLM as an arbiter or by having a human expert select the most realistic trajectory.

In conclusion, DT-GPT serves as a digital twin forecasting platform, enabling accurate and stable predictions, exploratory interpretability via a natural-language interface, and forecasting of patient variables not used in fine-tuning. DT-GPT exhibits true digital twin behaviors, potentially reproducing all aspects of the patients it represents, and surpassing traditional AI methods optimized for individual variables. We believe patient-level digital twins will impact clinical trials by supporting biomarker exploration, trial design, and interim analysis. Additionally, digital twins will assist doctors in treatment selection and patient monitoring. Overall, we envision LLM-powered digital twins becoming integral to healthcare systems.

## Data Availability

The Flatiron Health datasets are available upon request for the specific purpose of replicating results at. The MIMIC-IV dataset is publicly available.

## Acknowledgements

We would like to thank Anton Kraxner for providing crucial insights into NSCLC, as well as Ginte Kutkaite, Hugo Loureiro, Franziska Braun, Rudolf Kinder and Venus So for their valuable input and discussions.

## Source code and data access

The Flatiron Health data that support the findings of this study were originated by and are the property of Flatiron Health, Inc., which has restrictions prohibiting the authors from making the data set publicly available. Requests for data sharing by license or by permission for the specific purpose of replicating results in this manuscript can be submitted to. The Medical Information Mart for Intensive Care IV (MIMIC-IV) is available publicly online.^20^ The source code is in the process of being released, and will be shared openly via Github at publication time.

## Contribution statements

N Makarov and M Bordukova performed data processing, model implementation and model evaluation. R Rodriguez-Esteban, F Schmich and MP Menden supervised, designed and directed the project. All authors contributed to the conceptualisation and writing of the manuscript. N Makarov and M Bordukova had access to the underlying data, and all authors had access to the study data.

## Appendix

## A1 Dataset details

**Table A1.1:** Dataset details.

|  | Flatiron Health - NSCLC | ICU - MIMIC-IV |
| --- | --- | --- |
| # Patients | 16496 | 35131 |
| # Time Points | 773607 | 1686288 |
| # Input Variables | 320 | 300 |
| # Output Variables | 6 | 3 |
| Avg. % Missing in Input | 94.4% | 98.1% |
| Avg. % Missing in Output | 74.5% | 35.1% |
| Female/Male/NA % | 51.0/49.0/0.0 | 39.7/51.1/9.2 |
| Avg./Std. Age at Start | 67.5/10.2 | 64.0/16.3 |
| Avg./Std. Length of Full Patient Trajectory | 160.5/328.6 days | 46.78/0.87 hours |
| Avg./Std. Nr of Total Events per Patient Trajectory | 35.4/33.1 | 47.3/2.3 |
| Avg./Std. average length between events | 5.8/17.0 weeks | 1.0/0.2 hours |
| Time Point Resolution | Weekly | Hourly |
| Input Time Horizon | Unlimited | 24 hours |
| Forecast Time Horizon | Up to 13 weeks | Up to 24 hours |

The outlier processing method is outlined in **Appendix A2**. The splitting into training/validation/test datasets is performed randomly for the MIMIC-IV dataset, whilst stratified by group stage, smoking status, number of observations per visit and number of visits with drug administrations for the NSCLC dataset to ensure a balanced evaluation.

For the NSCLC dataset, we selected the number of laboratory variables to incorporate all variables that were already used in linear prognostic models [21], as well to have variables seen in at least 2000 patients. The number of diagnoses was chosen to include key information, as well as to have enough patient data for useful model training, having at least 1700 observations. With the clinical importance of variables shown in **Table A1.2**.

For the MIMIC-IV dataset, we define high variability as measured by the R^2^ for the last observed value for the full forecast. The lower the R^2^, the higher the variability.

**Table A1.2:** Details on the NSCLC output variables.

| Variable | LOINC | Impact |
| --- | --- | --- |
| Leukocytes [$10^9/L$] | 26464-8 | NSCLC treatment, particularly chemotherapy, can cause leukopenia, leading to decreased leukocyte counts. This reduction in leukocytes can increase the risk of infections due to a compromised immune system. |
| Lymphocytes/Leukocytes [%] | 26478-8 | This ratio is often used to monitor the immune status and inflammatory response. |
| Neutrophils [$10^9/L$] | 26499-4 | Chemotherapy can lead to neutropenia, resulting in a reduced neutrophil count. Neutropenia increases the risk of infections. |
| Lymphocytes [$10^9/L$] | 26474-7 | Lymphocyte counts often decrease during NSCLC treatment due to the immunosuppressive effects of chemotherapy. This reduction can impair the body's ability to fight infections and may affect the overall immune response. |
| Lactate Dehydrogenase [U/L] | 2532-0 | Elevated levels of lactate dehydrogenase (LDH) can be observed, indicating tissue damage or tumor burden. |
| Hemoglobin [g/dL] | 718-7 | Hemoglobin levels may decrease, leading to anemia, as a side effect of chemotherapy or due to the cancer itself. Anemia can cause symptoms such as fatigue, weakness, and shortness of breath, impacting the patient's quality of life. |

## A2 Outlier processing

To remove outliers, all target values more than three standard deviations were initially filtered out. Since there was still too much noise in the data, a second round of the filter was applied, though this time values were clipped, since high values could be outliers but still provide useful information.

## A3 Base pretrained LLM

We focus on biomedical LLMs as base LLMs, since they have been shown to contain biomedical knowledge, potentially improving the performance of the overall model. A number of different biomedical LLMs have been proposed, one of them BioMistral 7B DARE.^22^ We selected this model because it has an open source license, is based on a set of popular existing LLMs and has been shown to perform well on biomedical tasks. Additionally, following Gruver et. al, the model tokenizes numbers into individual digits.^16^ Here we use the 7B parameter version since it allows for a larger amount of experimentation with limited computational costs. Because the method is agnostic to the underlying LLM, DT-GPT can be equally developed based on larger models as well, which could potentially produce better forecasts.

## A4 Forecasting prompt examples

We structure the template in four components:

1. The patient’s history is noted down chronologically, using relative dating to prevent overfitting on time or date. For each patient visit and for each observed value, we note down the variable’s name and value, whilst omitting any missing variables.
2. Next, we include the patient’s baseline data, such as age and cancer stage
3. Since we do not impute target values, we include information about which variables should be in the output at which future time points.
4. Finally, we add a short prompt.

The target variables are also converted based on templates, containing only the respective target values. To reduce the amount of tokens required, the output is formatted so the target variable is provided followed by the list of values corresponding to the days that we want to output.

Note that for the MIMIC dataset, for each variable, if the value was observed in the patient’s history, it will be forward propagated, ensuring that we have the information even if the context length is normally not long enough. Here, we present synthetic examples of both the manual template and JSON input as well as output.

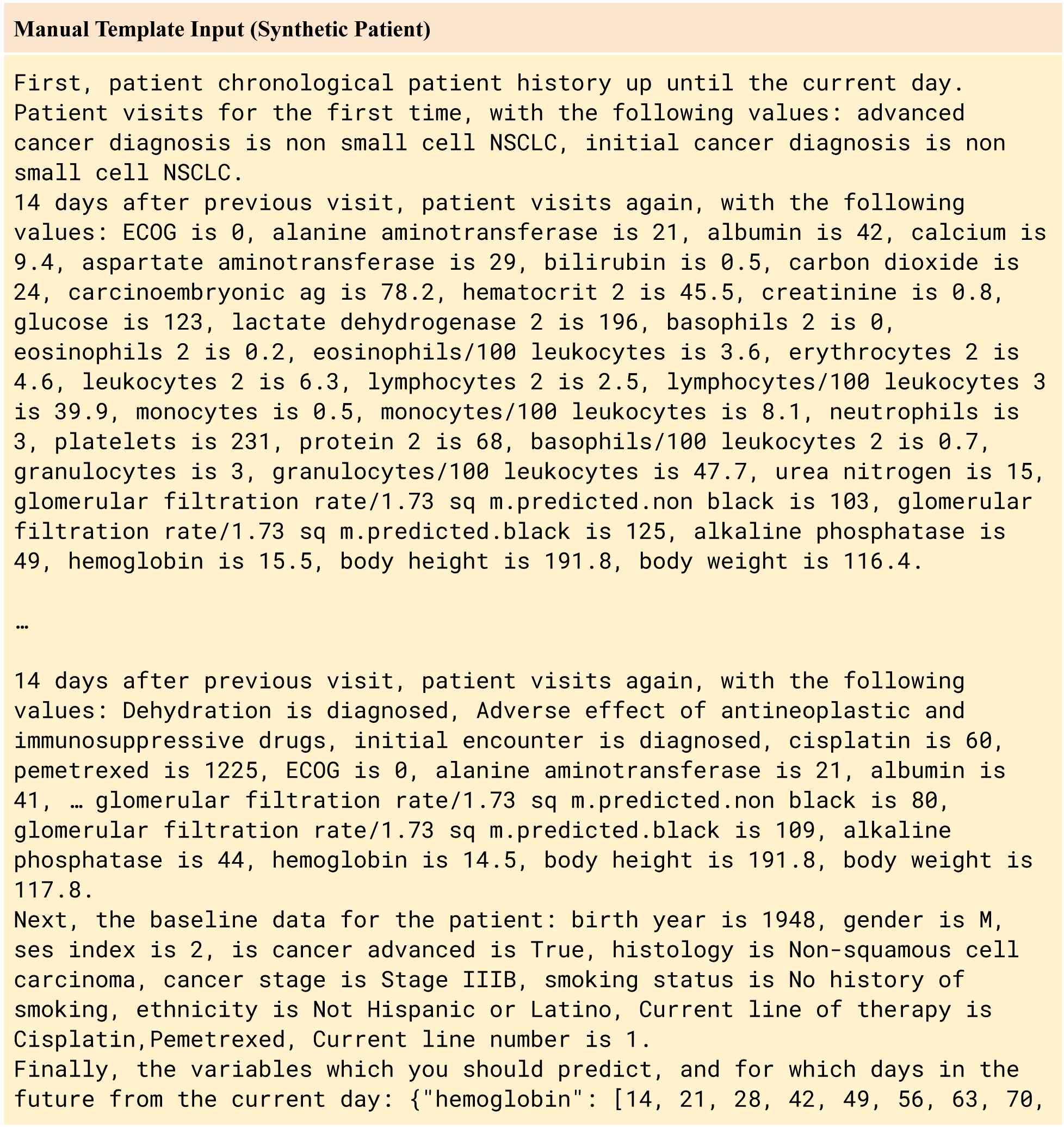
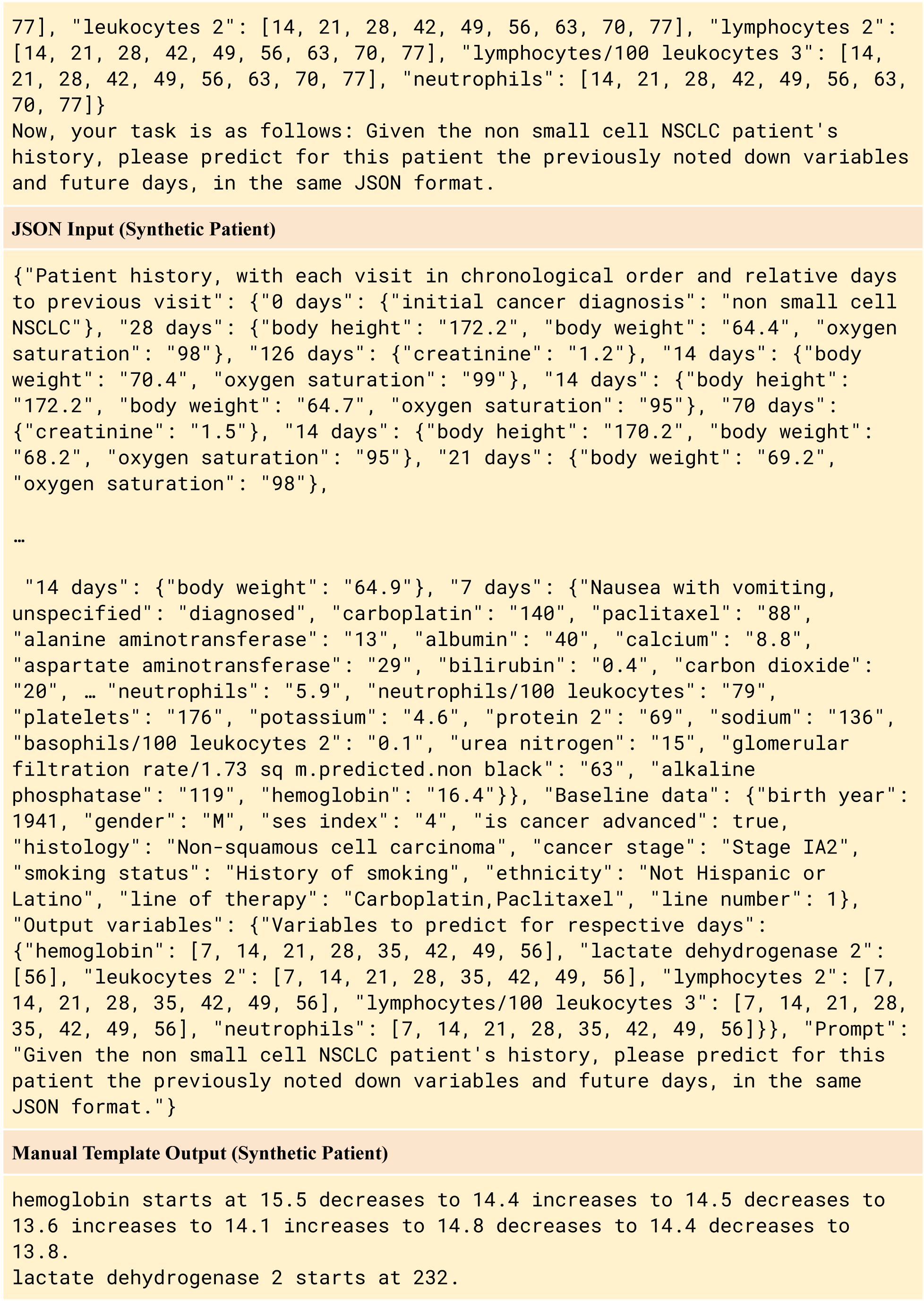
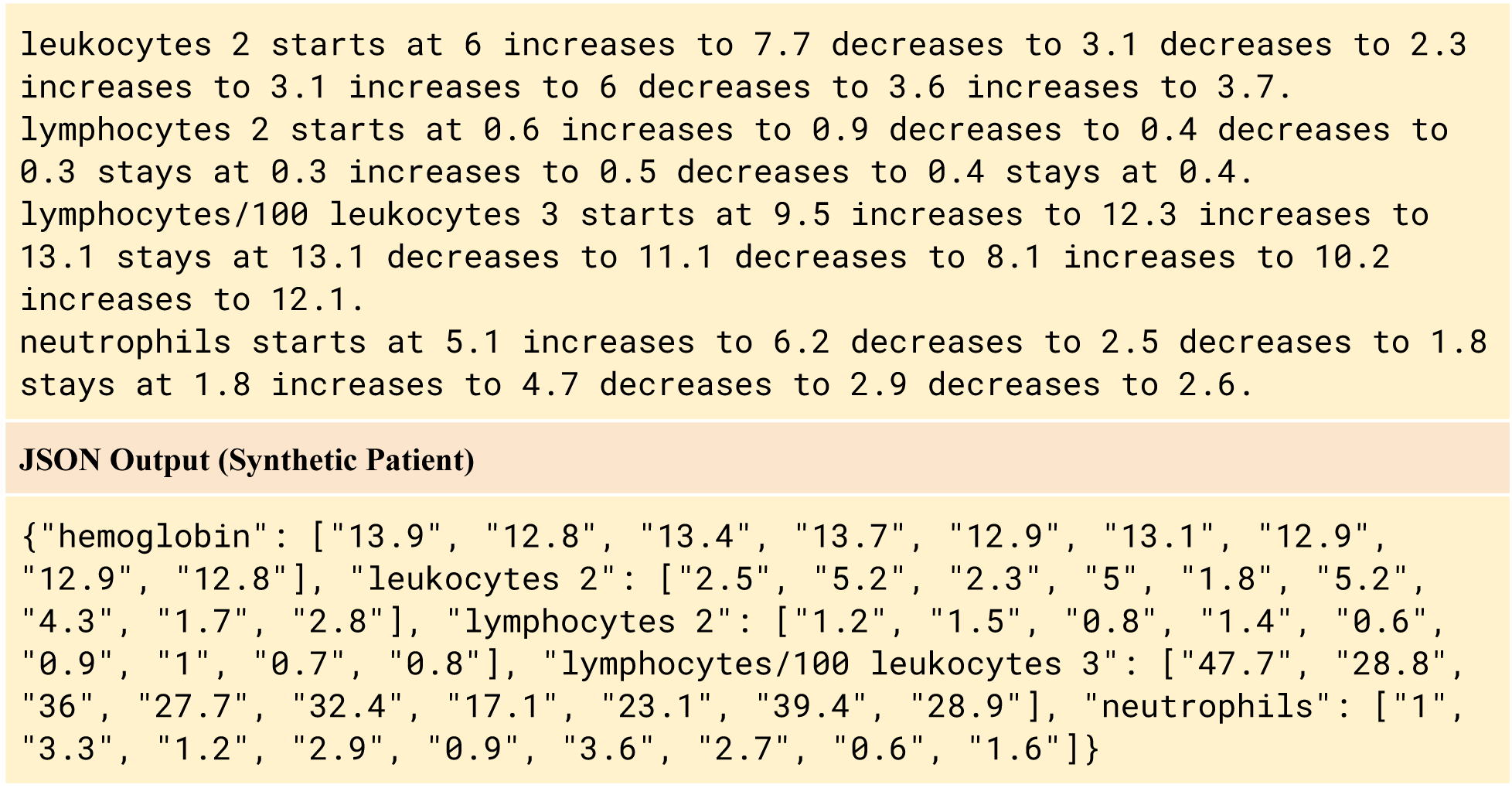

## A5 Fine-tuning & inference details

We initially experimented with applying the loss to the entire sequence, which would also allow generating synthetic patients, however the models hallucinated to an usable point. Instead we employed a masking such that the gradient is only computed for the tokens that need to be forecast. For the training, we set the learning rate to 10^−5^, a warm up ratio of 0·1, batch size of 1, employ a cosine learning rate scheduler, with a weight decay 0·1 and the optimizer being AdamW. During training, we limit the input sequence length to 3400 tokens due to memory constraints. The optimal epoch was identified based on the loss on the validation set, with the training taking around 20 hours on a single NVIDIA A100 80GB GPU. For all evaluations, we run the model 30 times on each patient sample, and a maximum final sequence length of 4000 tokens. We used nucleus sampling with top p set to 0·9 and temperature set to 1·0.

For the chatbot prediction explainability and zero-shot non-target variable forecasting, we used the same nucleus sampling parameters (top p = 0·9 and temperature = 1). The maximum sequence length was set to 200 tokens for the explainability task and 120 tokens for the zero-shot forecasting task, respectively. The numbers were selected to cover the desired output sequence length and prevent hallucinations. For the zero-shot forecasting, we run DT-GPT 10 times on each patient sample and use mean aggregation to obtain the final prediction.

In the context of patient digital twins, it is crucial to differentiate between simulation and forecasting. Simulations represent realistic patient trajectories, whereas forecasts predict the trajectories that are most likely to happen. Ideally, simulations should be able to cover the distribution of all possible patient trajectories.

## A6 Chatbot prompt examples

A two-step chatbot interaction example for the prediction explainability task is provided below.

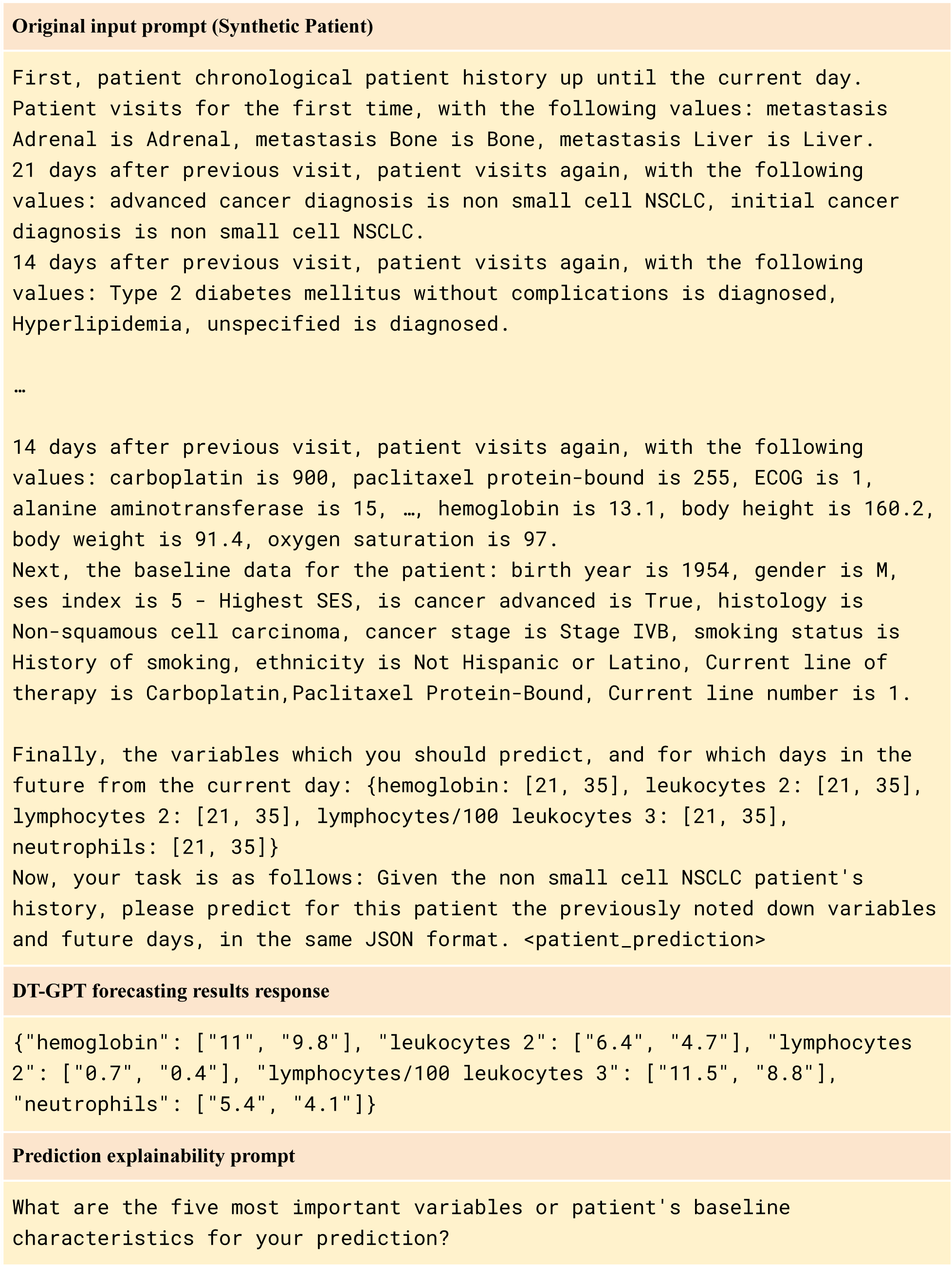
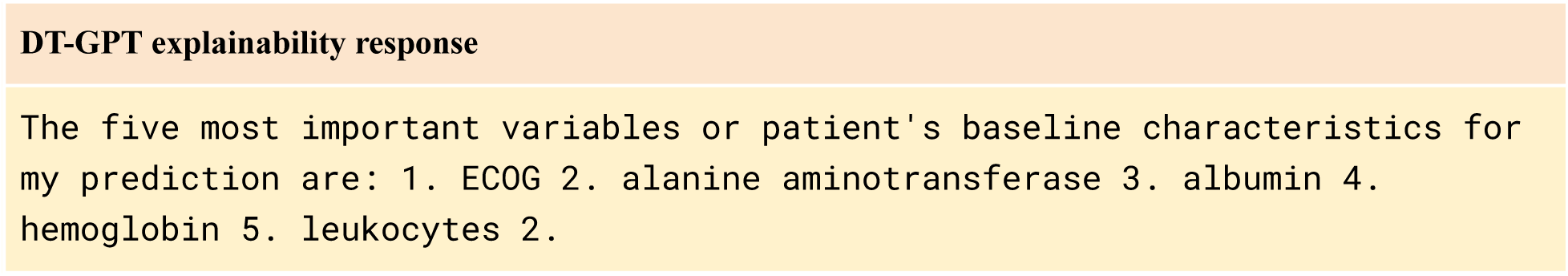

A two-step chatbot interaction example for the zero-shot forecasting task is provided below.

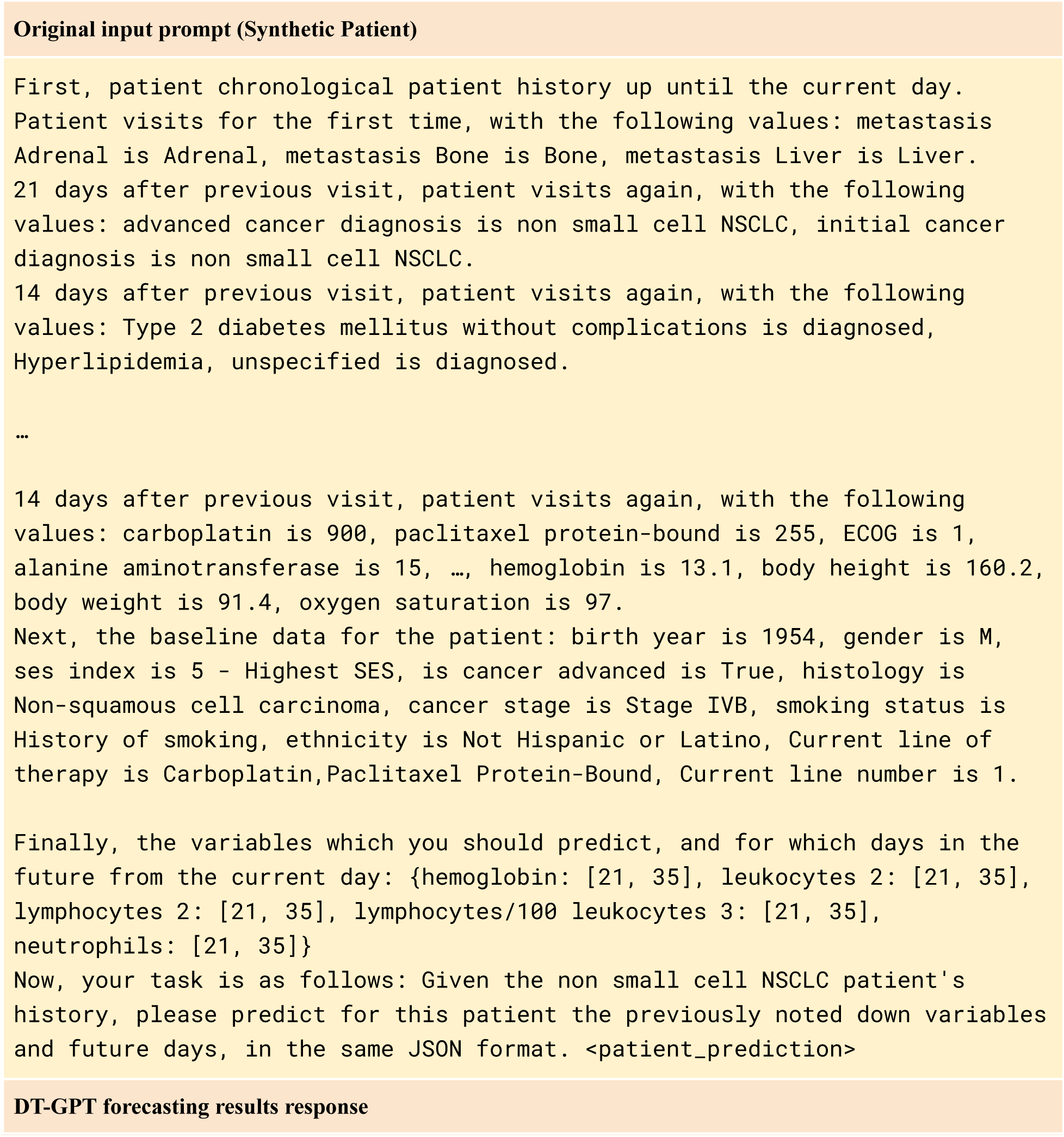
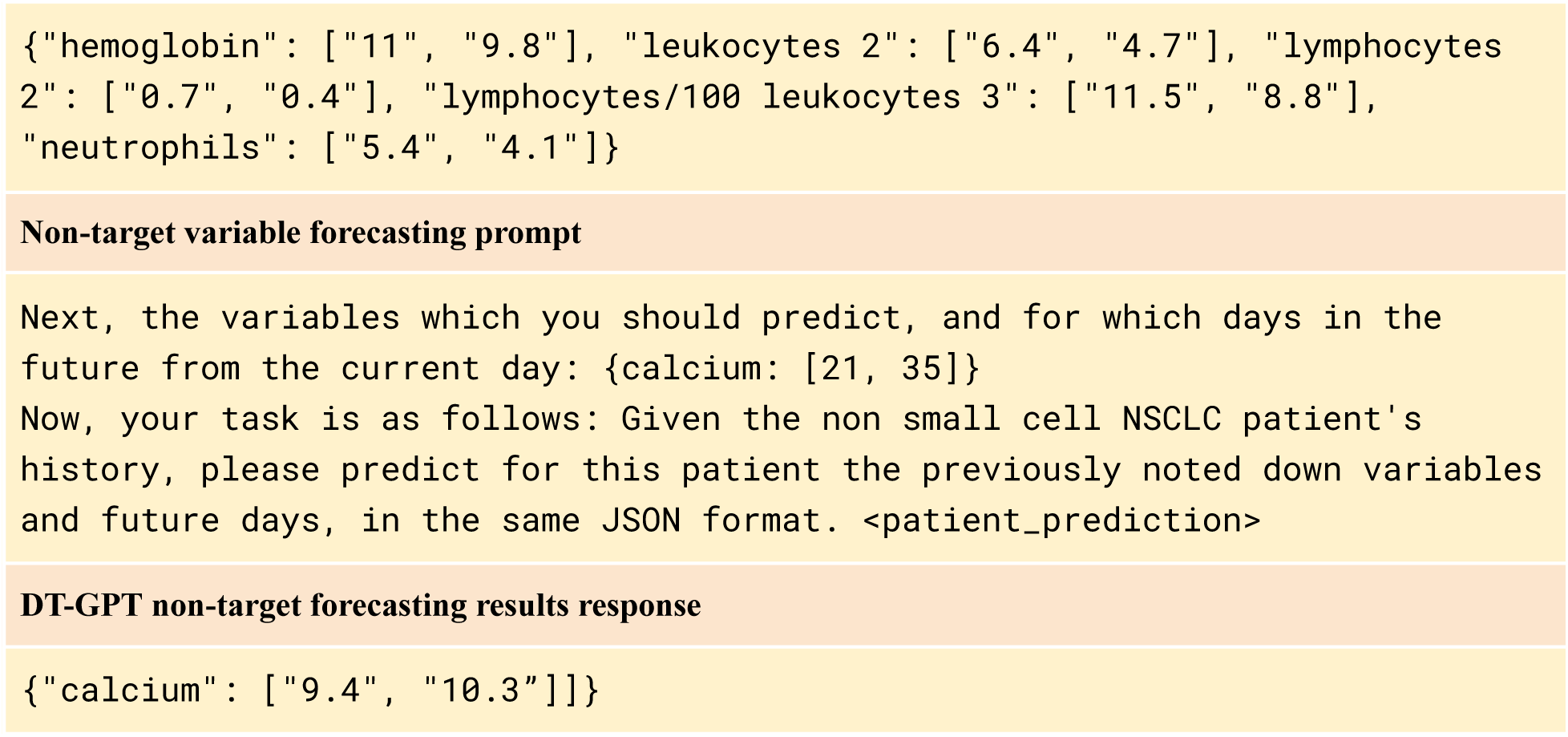

## A7 Baselines & metric details

The baseline models are implemented in the Darts library and the default hyperparameters are used. For the input time horizon, both 35 and 91 days were explored for the Flatiron Health NSCLC dataset, whilst the full 24 hours was used for the MIMIC dataset. Since the models cannot natively deal with missing data, we employ linear interpolation with forward and backward passes on the input data, and linear interpolation only with forward pass on the target data. We apply the filtering based on three standard deviations, as with DT-GPT, and then apply standardization or one hot encoding. To ensure fairness between the baseline models and DT-GPT, we also provide the baselines with an indicator variable, having 1 for every future date which will be measured and 0 for those which are imputed.

## A8 Results

In **Tables A8.1** and **A8.2** we show the performance of the models across the two datasets. It is interesting to note that LightGBM performs better than more complex models, which we hypothesize is due to the high dimensional noisy data, though this has also been observed in the literature.^26,27^

**Table A8.1:** Performance of all models on the NSCLC dataset. “Corr.” means Spearman Correlation (higher is better), “MAE” is Mean Absolute Error (lower is better), with the best performance highlighted in bold and ranked by the average MAE.

|  | NSCLC |  |  |  |  |  |  |  |  |  |  |  |
| --- | --- | --- | --- | --- | --- | --- | --- | --- | --- | --- | --- | --- |
| Model | Hemoglobin |  | Leukocytes |  | Lymphocytes/Leukocytes |  | Lymphocytes |  | Neutrophils |  | Lactate Dehydrogenase |  |
|  | MAE | Corr. | MAE | Corr. | MAE | Corr. | MAE | Corr. | MAE | Corr. | MAE | Corr. |
| Copy Forward | 0.698 | 0.629 | 0.969 | 0.333 | 0.731 | 0.499 | 0.569 | 0.603 | 0.974 | 0.323 | 0.433 | **0.739** |
| Linear Regr. | 0.486 | 0.759 | 0.782 | 0.441 | 0.668 | 0.545 | 0.506 | 0.656 | 0.778 | 0.410 | 0.475 | 0.683 |
| TFT | 0.469 | 0.774 | 0.719 | 0.504 | 0.651 | 0.563 | 0.463 | 0.696 | 0.717 | 0.476 | 0.480 | 0.646 |
| TiDE | 0.464 | 0.768 | 0.737 | 0.488 | 0.655 | 0.567 | 0.465 | 0.704 | 0.740 | 0.452 | 0.453 | 0.668 |
| Light GBM | 0.453 | 0.780 | 0.727 | 0.508 | **0.644** | 0.583 | 0.456 | 0.712 | 0.734 | 0.467 | 0.425 | 0.732 |
| DT-GPT (ours) | **0.440** | **0.796** | **0.689** | **0.540** | 0.650 | **0.587** | **0.437** | **0.733** | **0.699** | **0.496** | **0.417** | 0.731 |

**Table A8.2:** Performance of all models on the ICU dataset. “Corr.” means Spearman Correlation (higher is better), “MAE” is Mean Absolute Error (lower is better), with the best performance highlighted in bold and ranked by the average MAE.

|  | Intensive Care Unit |  |  |  |  |  |
| --- | --- | --- | --- | --- | --- | --- |
| Model | Magnesium |  | Respiratory Rate |  | O2 Saturation |  |
|  | MAE | Corr. | MAE | Corr. | MAE | Corr. |
| Copy Forward | 0.681 | 0.462 | 0.769 | 0.470 | 0.746 | 0.484 |
| Linear Regression | 0.606 | 0.463 | 0.680 | 0.509 | 0.681 | 0.525 |
| TiDE | 0.534 | 0.549 | 0.635 | 0.559 | 0.652 | 0.570 |
| Temporal Fusion Transformer | 0.537 | 0.555 | 0.635 | **0.562** | 0.644 | **0.576** |
| LightGBM | 0.520 | 0.583 | **0.634** | **0.562** | 0.644 | 0.573 |
| DT-GPT (ours) | **0.505** | **0.609** | 0.636 | **0.562** | **0.635** | **0.576** |

**Figure A8.1:**
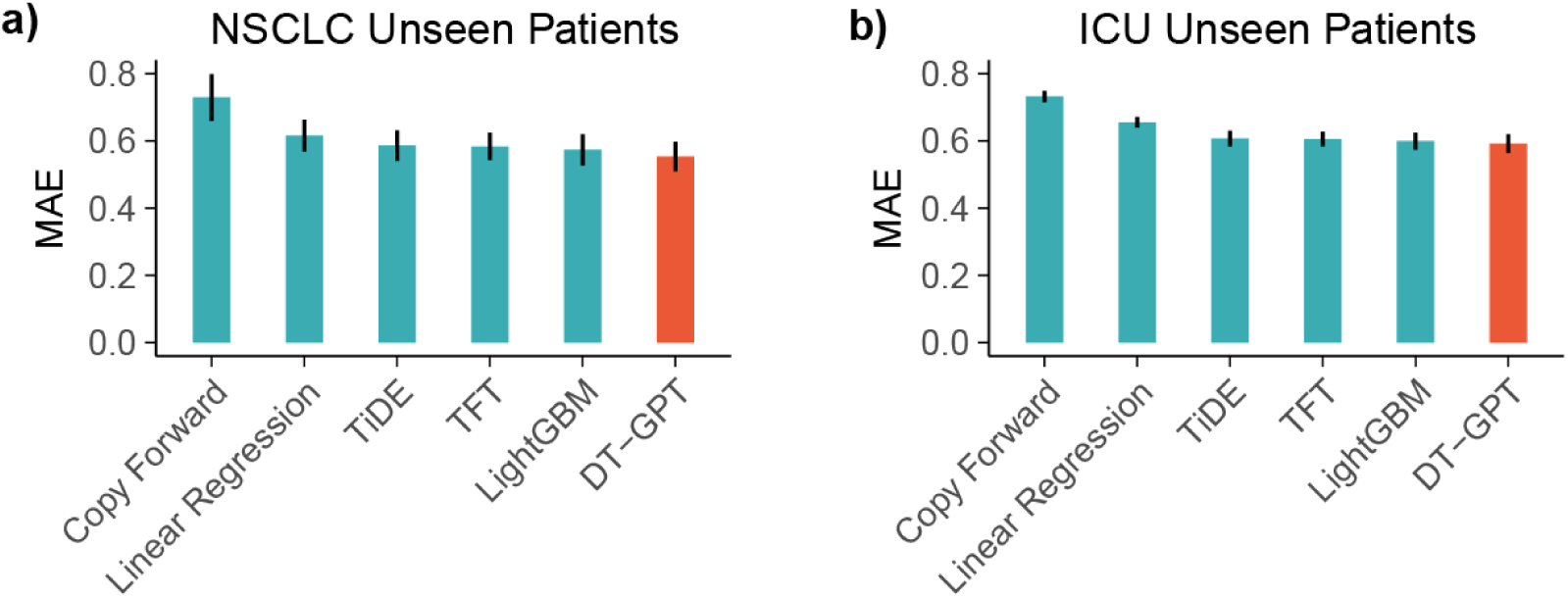
**a)** and **b)** show a comparison of all baseline models on the NSCLC and ICU datasets, respectively, with error bars showing the standard error across all variables.

## A9 Perturbation study details

The misspelling algorithm randomly performs either perturbation, insertion, deletion or replacement, using all ASCII letters & digits, applied to the entire input text. This includes dates, variable names, values, baseline information and prompts. One operation is considered one misspelling.

## A10 Encoding method comparison

**Figure A10.1:**
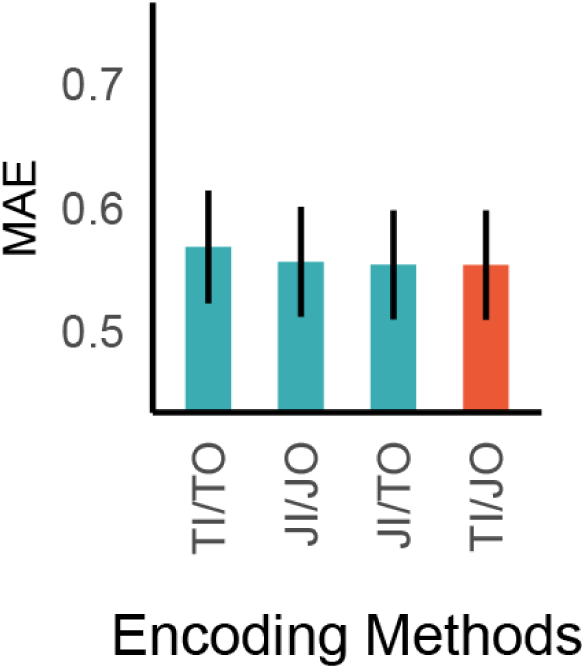
Different encoding methods and their respective performances on mean absolute error (MAE), with the following abbreviations: TI - “Text Input”, TO - “Text Output”, JI - “JSON Input” and JO - “JSON Output”.

DT-GPT is stable with respect to different data encoding strategies (**Fig. A10.1)**, though Text In, JSON Out (TI/JO) and JSON In, TEXT Out (JI/TO) perform best, with TI/JO being marginally more efficient. Specifically Text In, Text Out (TI/TO) achieves an average MAE of 0·568 ± 0·05, JSON In, JSON Out (JI/JO) reaches 0·556 ± 0·04, JSON In, JI/TO reaches 0·554 ± 0·04, TI/JO attains 0·554 ± 0·04.

## A11 Most important variables and patient baseline characteristics for the forecasting

For each of 2,773 patient samples in the test set sample, we obtain 10 predicted trajectories and 5 variables or patient baseline characteristics explaining those trajectories.We present the percentage of trajectories explained by the most important variables in the table below (**Tab. A11.1)**.

**Table A11.1:** Key variables and the percentage of predicted trajectories they explain as extracted from DT-GPT.

| Variable | % of predicted trajectories it explains according to DT-GPT |
| --- | --- |
| therapy | 87.0 |
| ECOG | 55.6 |
| leukocytes | 44.5 |
| age | 36.2 |
| alanine aminotransferase | 26.0 |
| hemoglobin | 21.8 |
| neutrophils | 21.3 |
| lymphocytes/100 leukocytes | 21.0 |
| body weight and body height | 19.4 |
| body height | 19.4 |
| albumin | 16.5 |
| gender | 15.8 |
| lymphocytes | 15.8 |
| lactate dehydrogenase 2 | 12.3 |
| alkaline phosphatase | 9.5 |
| ferritin | 7.2 |

## A12 Performance results of zero-shot DT-GPT and LightGBM baselines for non-target forecasting task

We visualize and investigate the influence of the five most important variables, therapy (**Fig. A12.1**), ECOG (**Fig. A12.2**), age (**Fig A12.3**), leukocytes (**Fig A12.4**) and alanine aminotransferase (**Fig A12.5**) on the dynamics of predicted NSCLC output variables. While the variables were marked as important for the prediction of six output variables simultaneously, we note that most of them particularly influence dynamics in only a subset of output variables. For instance, therapy is correlated with neutrophils, hemoglobin and leukocytes trajectories (**Fig. A12.1**), ECOG with hemoglobin and lymphocytes to leukocytes ratio trajectories (**Fig. A12.2**), and age with lactate dehydrogenase (**Fig A12.3**), respectively.

For the therapy, we consider 10 most frequent therapies and group them into therapy group as follows: carboplatin & paclitaxel, carboplatin & pemetrexed and pemetrexed for the chemotherapy group (450 patients), pembrolizumab, nivolumab and durvalumab for the immunotherapy group (598 patients), carboplatin & pembrolizumab & pemetrexed, docetaxel & ramucirumab, pembrolizumab & pemetrexed for the combination therapy group (434 patients) and osimertinib for the target therapy group (112 patients), respectively. For the ECOG variable, we consider values of 0 (765 patients), 1 (1299 patients), 2 (384 patients) and 3 (85 patients). For the age, we define the following groups based on the age histogram: younger than 50 years old (124 patients), between 50 and 60 years old (474 patients), between 60 and 70 years old (911 patients), between 70 and 80 years old (981 patients) and older than 80 years old (261 patients). For leukocytes, we consider the last observed value within 13 weeks of medical history prior to the start of the treatment as a baseline value, and based on the histogram of leukocytes baseline values combined with the reference intervals for the leukocytes define four groups: less than 5 10^9^/L (315 patients), between 5 and 10 10^9^/L (1093 patients), between 10 and 20 10^9^/L (488 patients) and more than 20 10^9^/L (66 patients). Similarly, for the alanine aminotransferase (ALT) we consider the last observed value within 13 weeks of medical history prior to the start of the treatment as a baseline value, and based on the histogram of ALT baseline values combined with the reference intervals for the ALT define four groups: less than 10 U/L (365 patients), between 10 and 20 U/L (1090 patients), between 20 and 30 U/L (602 patients), between 30 and 40 U/L (249 patients) and more than 40 U/L (246 patients). For each of the most important variables, only predicted trajectories of patients with available variable values and ground truth for output variables were analyzed.

Since we ask DT-GPT to explain all output variables simultaneously, the observed important variables do not lead to a “perfect” separation of predicted trajectories conditioned on the variable value. For instance, if we perform forecasting of hemoglobin trajectories only, further variables are said to be important by DT-GPT (**Appendix A13**).

Furthermore, the choice of groups in the following analysis can be made arbitrary and might influence the results. A better grouping approach would include other variables such as age, gender or other demographics or laboratory test data. Specifically, hemoglobin and ALT have different reference intervals for males and females. Such interactions were outside of the scope of this analysis. Finally, we note that we can establish only the correlation between obtained important variables and predicted trajectories; the causal relationships are more complex and are subject to further investigation.

**Figure A12.1:**
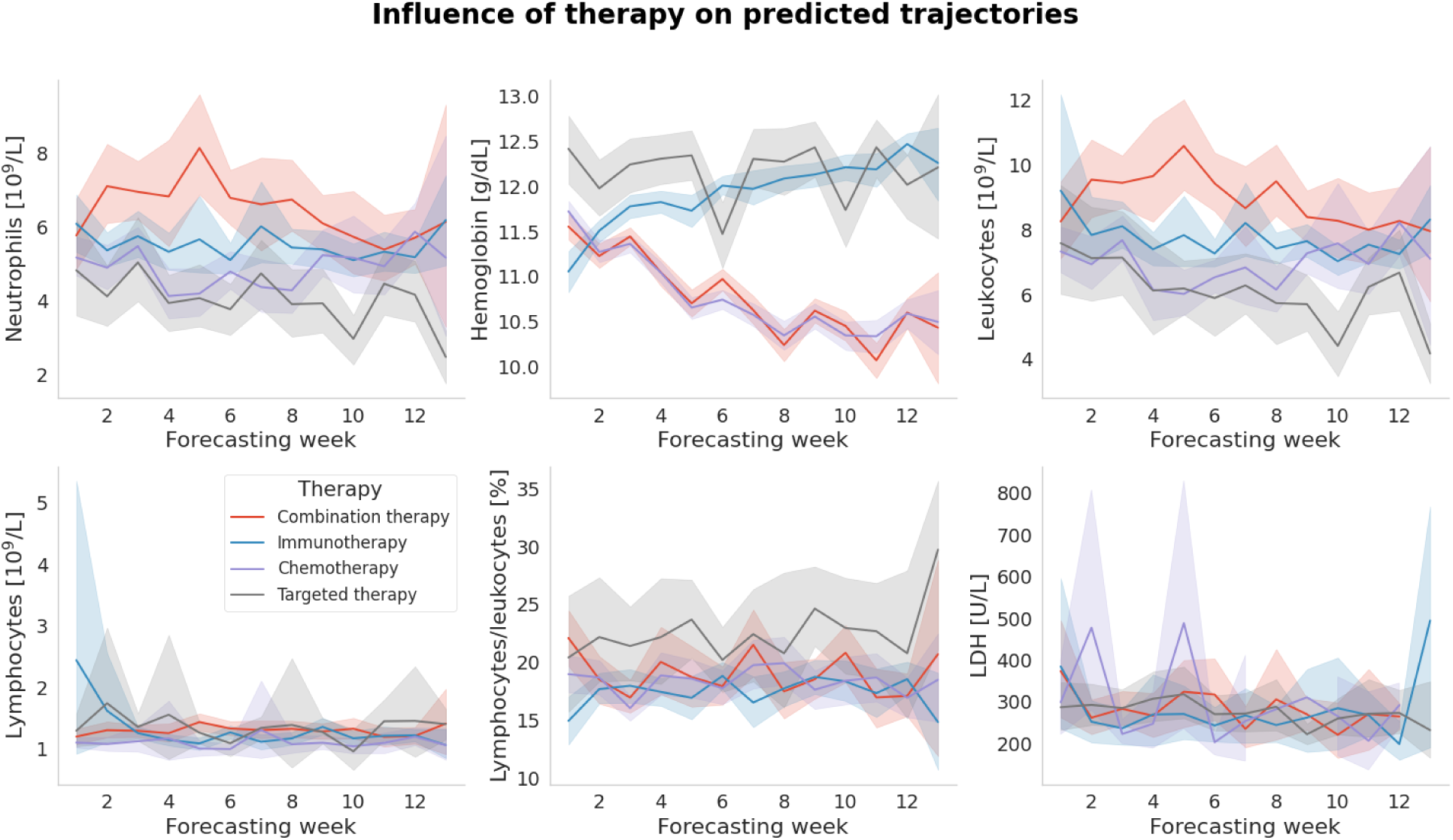
The most important variable, therapy particularly influences the predicted dynamics of neutrophils, hemoglobin, leukocytes and lymphocytes to leukocytes ratio. Here, lines represent average trajectories with 95% confidence intervals calculated through bootstrapping.

**Figure A12.2:**
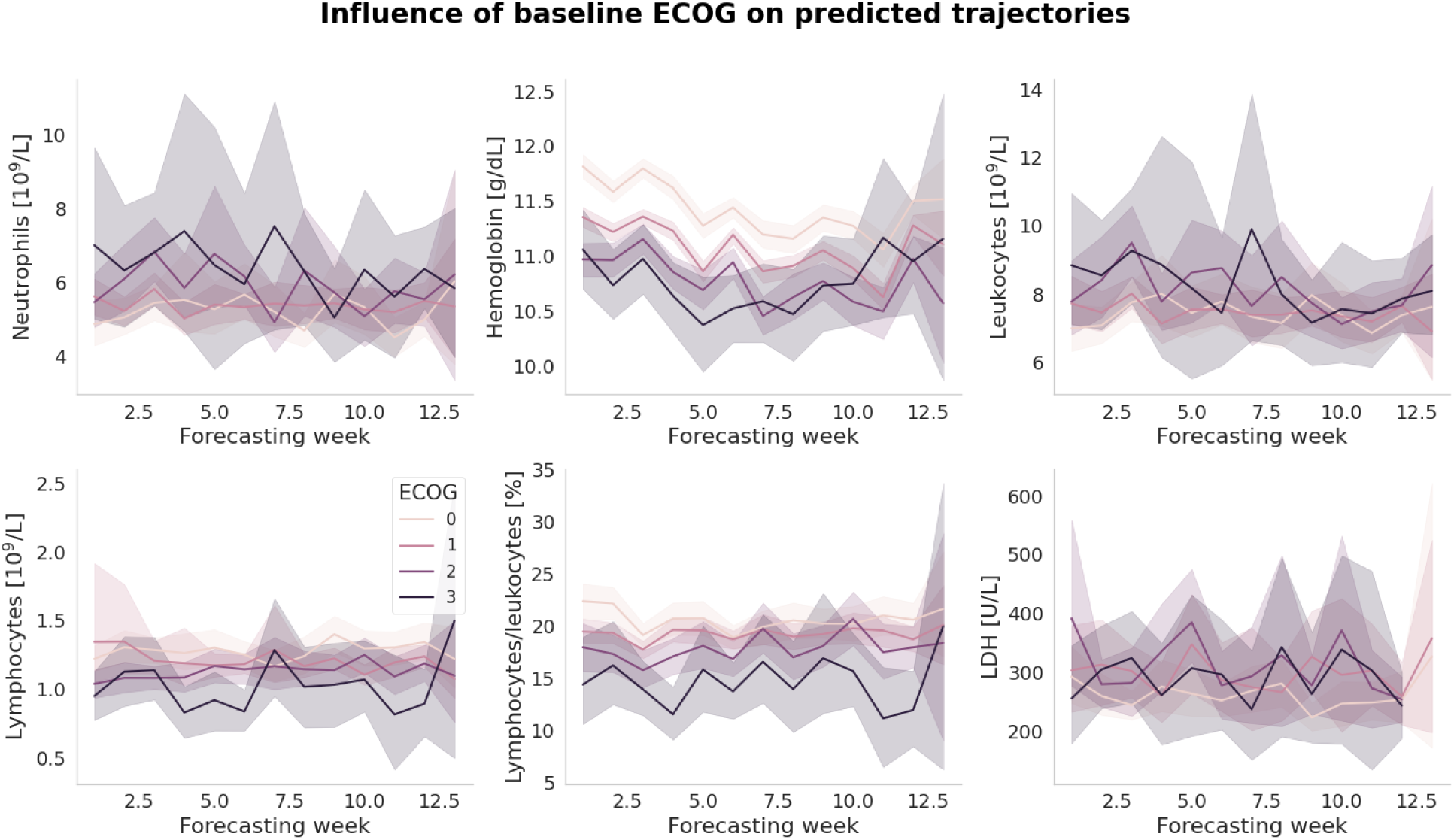
The second most important variable, ECOG, particularly influences the predicted dynamics of hemoglobin and lymphocytes to leukocytes ratio. Here, lines represent average trajectories with 95% confidence intervals calculated through bootstrapping.

**Figure A12.3:**
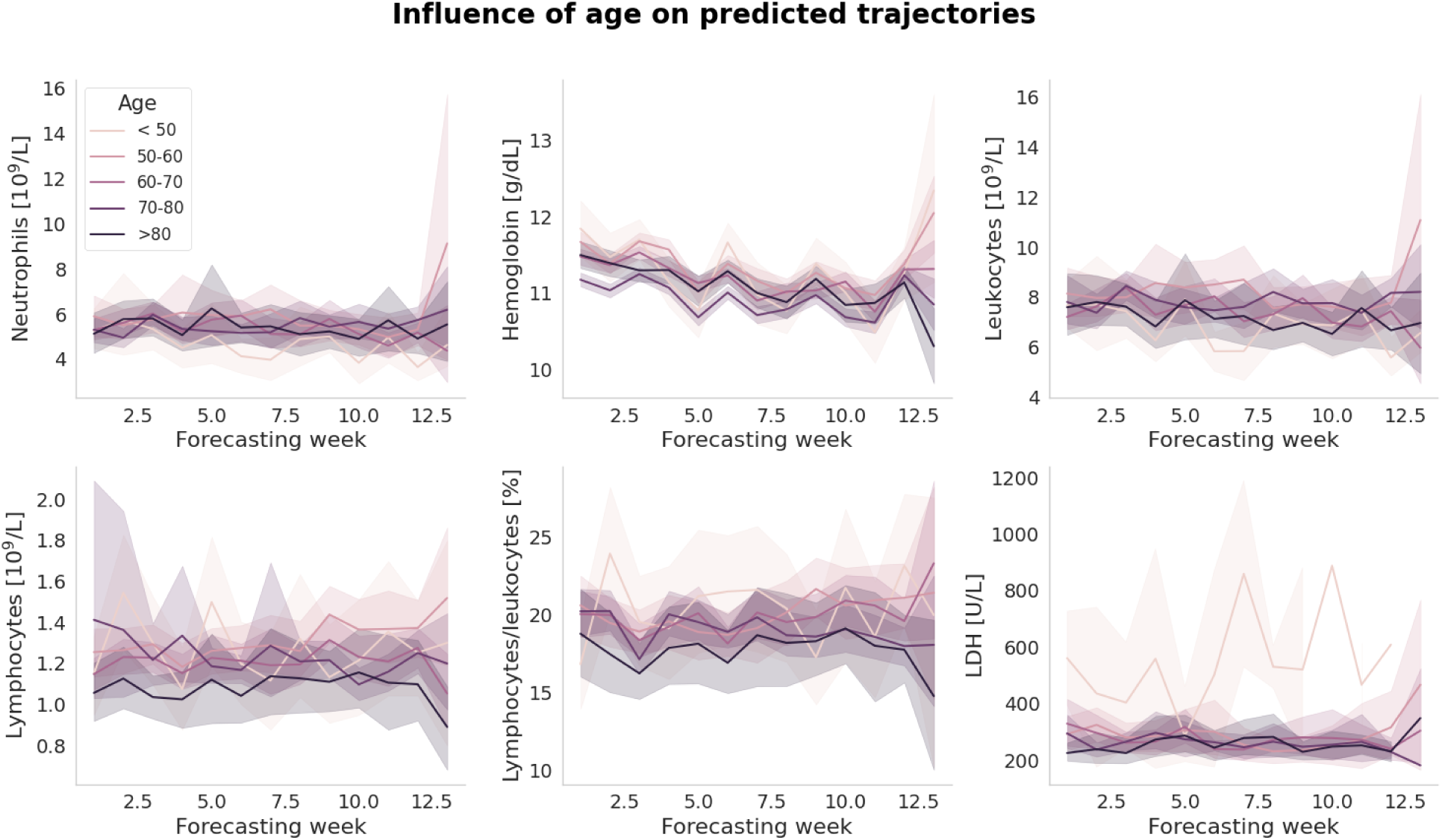
The third most important variable, age, particularly influences the predicted dynamics of lactate dehydrogenase (LDH), whereby younger patients (less than 50 years old) have on average higher LDH values. Here, lines represent average trajectories with 95% confidence intervals calculated through bootstrapping.

**Figure A12.4:**
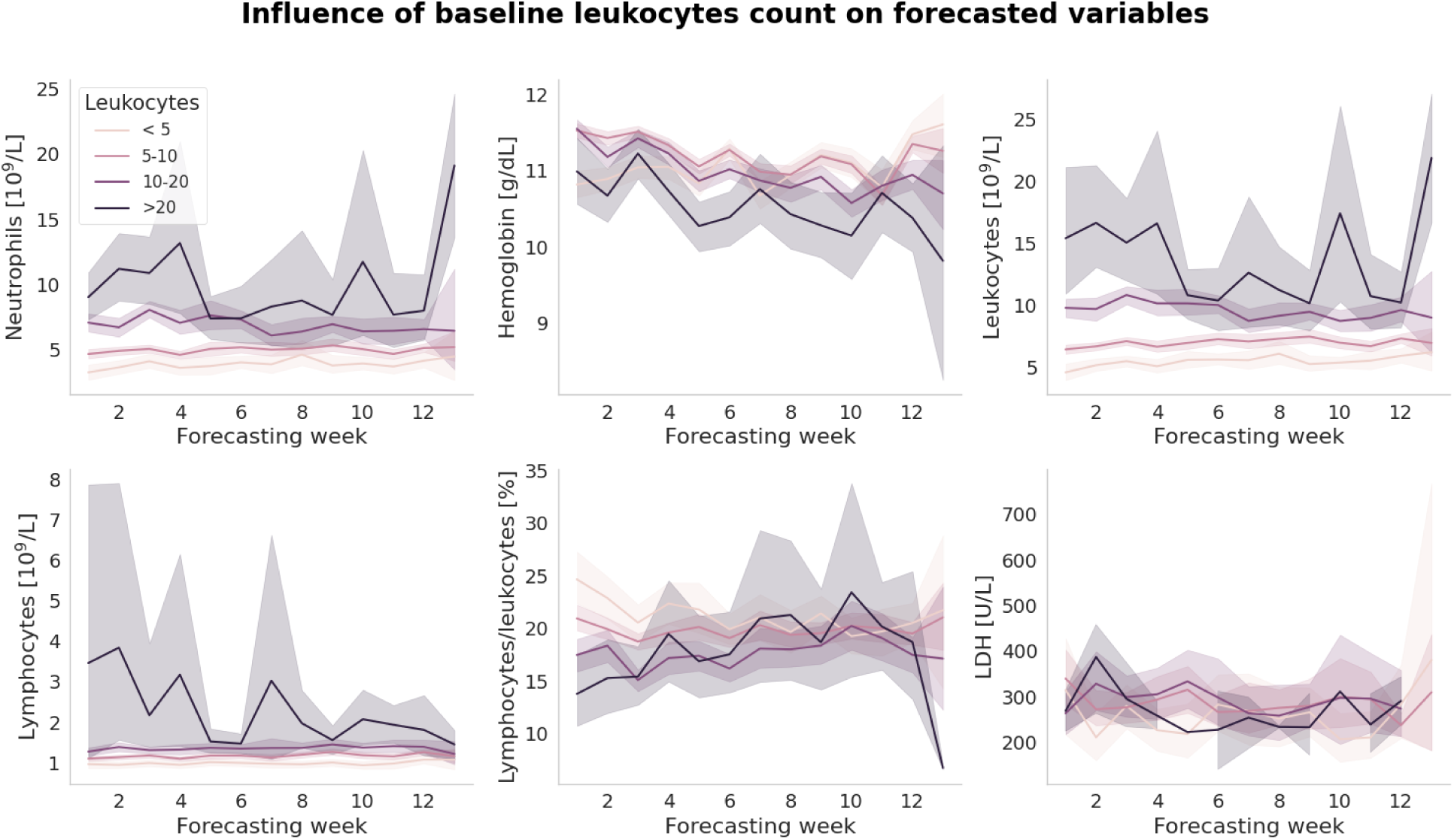
The fourth most important variable, leukocytes, particularly influences the predicted dynamics of neutrophils, hemoglobin, leukocytes and lymphocytes to leukocytes ratio. Here, lines represent average trajectories with 95% confidence intervals calculated through bootstrapping.

**Figure A12.5:**
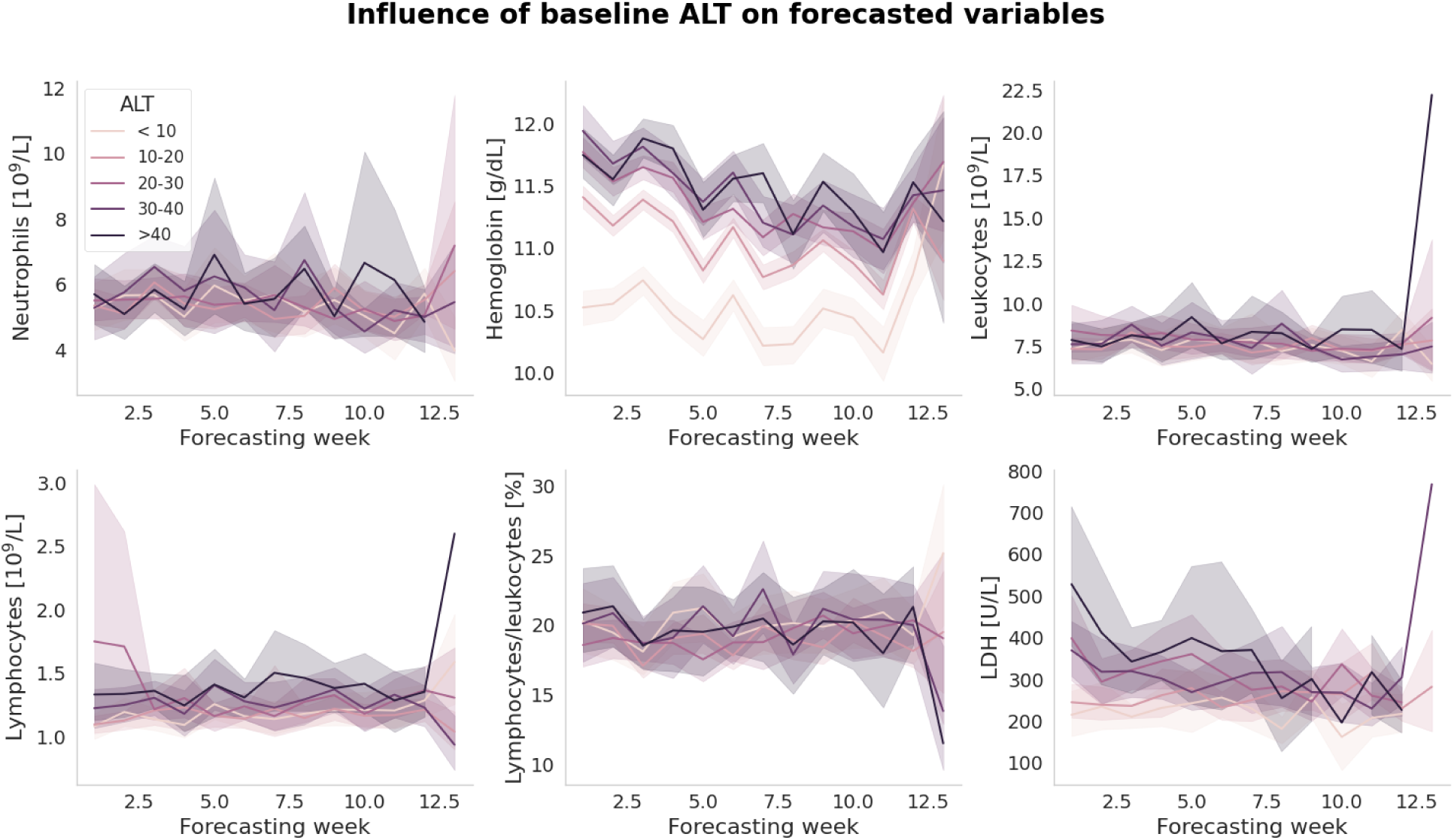
The fifth most important variable, alanine aminotransferase (ALT), particularly influences the predicted dynamics of neutrophils, hemoglobin. Here, lines represent average trajectories with 95% confidence intervals calculated through bootstrapping.

## A13 Explainability of predicted hemoglobin trajectories

We highlight complex and non-trivial explanatory abilities of DT-GPT on an example of hemoglobin trajectories. A set of obtained important variables for predicted hemoglobin trajectories is not constant across all trajectories, but is correlated with the predicted values. To show this, we first perform clustering of predicted hemoglobin trajectories as follows: (1) for each of the trajectories we calculate its mean value over time, (2) we consider distribution of mean values of trajectories and determine 0·15 and 0·75 quantiles of the distribution, (3) we assign trajectories with the mean value below 0·15 quantile to the low level group (mean value less than 9·76 g/dL), trajectories with the mean value above 0·85 quantile to the high level group (mean value greater than 13·31 g/dL), and trajectories with the mean value between 0·15 and 0·85 quintiles to the middle level group, respectively. For each of the groups we consider 10 most important variables separately and determine their intersection to define a final set of 16 important variables (**Fig A13.1**).

The fractions of important variables explaining hemoglobin trajectories (e.g., relative frequency of a variable to be in the set of important variables as outputted by DT-GPT for the trajectories in each group) are different for each of the hemoglobin group. While therapy, ECOG and age are the most frequent and considered as important for all three groups, some of the variables are more present in one of the groups, e.g., height for the high hemoglobin group or ALT and ferritin for the low level hemoglobin group (**Fig A13.1**). To quantify observed correlations statistically, we performed a chi-squared test for independence on the contingency table of the hemoglobin group versus the number of trajectories explained by each important variable. The null hypothesis of independence was rejected with p-value < 2.2e-16. Thus, DT-GPT might consider different variables when predicting hemoglobin trajectories of different levels highlighting its complex prediction explainability potential. However, our analysis only reveals correlations between hemoglobin level and important variables, thus causal claims are out of scope.

**Figure A13.1:**
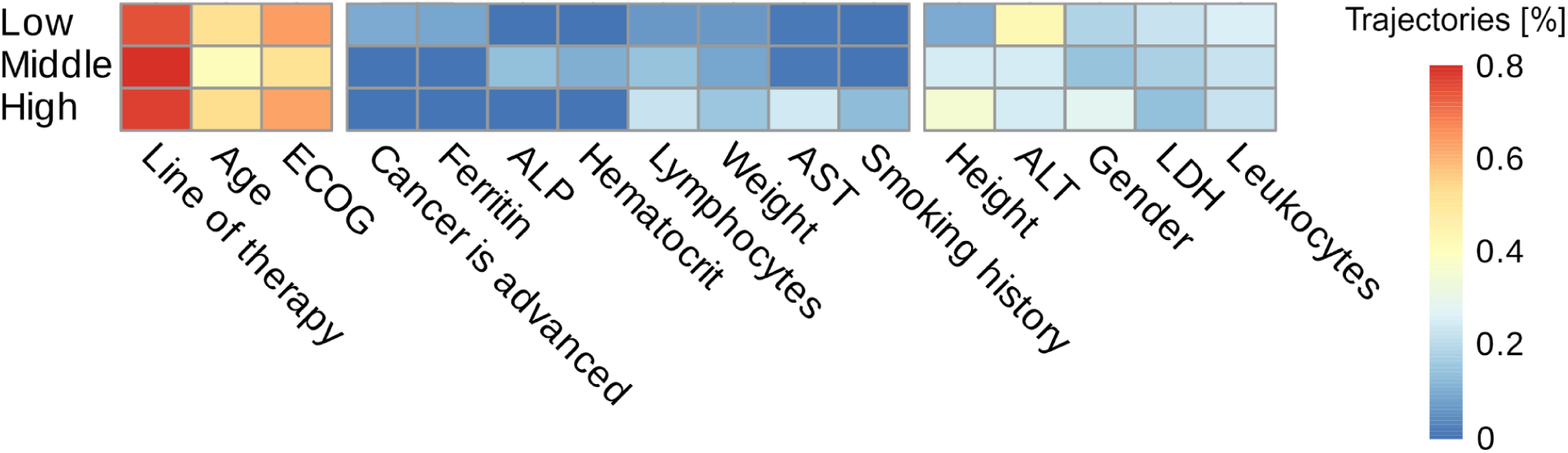
Predicted hemoglobin trajectories in low, middle and high level groups have different distributions of the important variables explaining them. While therapy, age and ECOG are present in each of the groups, the ferritin and “cancer is advanced” variables have a slightly higher prevalence in the lower group whilst gender is more frequent for the high hemoglobin level group. Chi-squared test for independence rejected the null hypothesis (p-value < 2·2e-16) indicating that DT-GPT might put more weight on different variables when predicting hemoglobin trajectories of different levels. (Abbreviations: ECOG - Eastern Cooperative Oncology Group performance status scale, LDH - Lactate dehydrogenase, ALT - Alanine aminotransferase, ALP - Alkaline phosphatase, AST - Aspartate aminotransferase).

## A14 Performance results of zero-shot DT-GPT and LightGBM baselines for non-target forecasting task

From the original 80 lab variables, we could evaluate the zero shot performance on 69 (**Tab. A14.1)**. The difference is due to the 6 variables used in training and a further 5 which had too few samples on either the input or output.

**Table A14.1.** Zero shot performance of DT-GPT on variables that were previously not trained.

| **Variable** | **LOINC** | **MAE**<br>**LightGBM** | **MAE**<br>**DT-GPT** |
| --- | --- | --- | --- |
| ferritin | 2276-4 | 0.07 | 0.03 |
| lactate dehydrogenase | 14804-9 | 0.55 | 0.14 |
| erythrocytes | 789-8 | 0.15 | 0.27 |
| carcinoembryonic ag | 2039-6 | 0.29 | 0.29 |
| erythrocytes 2 | 26453-1 | 0.26 | 0.37 |
| neutrophils.segmented | 30451-9 | 0.71 | 0.45 |
| lymphocytes 3 | 731-0 | 0.48 | 0.47 |
| neutrophils.band form | 26507-4 | 0.65 | 0.55 |
| neutrophils 2 | 751-8 | 0.70 | 0.58 |
| lymphocytes/100 leukocytes | 736-9 | 0.66 | 0.60 |
| lymphocytes | 732-8 | 0.68 | 0.60 |
| granulocytes | 30394-1 | 0.78 | 0.67 |
| leukocytes | 6690-2 | 0.71 | 0.69 |
| lymphocytes/100 leukocytes 2 | 737-7 | 0.75 | 0.70 |
| bilirubin.non-glucuronidated | 1971-1 | 0.57 | 0.71 |
| monocytes | 26484-6 | 0.72 | 0.71 |
| granulocytes 2 | 20482-6 | 0.76 | 0.72 |
| alkaline phosphatase | 6768-6 | 0.46 | 0.78 |
| coagulation tissue factor induced | 5902-2 | 0.58 | 0.81 |
| urea nitrogen | 3094-0 | 0.57 | 0.84 |

| Variable | LOINC | MAE<br>LightGBM | MAE<br>DT-GPT |
| --- | --- | --- | --- |
| glomerular filtration rate/1.73 sq m.predicted 2 | 69405-9 | 0.40 | 0.88 |
| protein | 2888-6 | 0.60 | 0.93 |
| monocytes/100 leukocytes 2 | 5905-5 | 0.78 | 0.93 |
| platelets 2 | 777-3 | 0.64 | 0.97 |
| glomerular filtration rate/1.73 sq m.predicted.black | 48643-1 | 0.39 | 1.05 |
| platelets | 26515-7 | 0.64 | 1.07 |
| hematocrit 2 | 20570-8 | 0.49 | 1.10 |
| glomerular filtration rate/1.73 sq m.predicted.non black | 48642-3 | 0.39 | 1.12 |
| calcium | 17861-6 | 0.64 | 1.14 |
| creatinine renal clearance.predicted | 35591-7 | 0.33 | 1.16 |
| potassium 2 | 6298-4 | 0.71 | 1.17 |
| coagulation tissue factor induced.inr | 38875-1 | 0.33 | 1.27 |
| urate | 3084-1 | 0.51 | 1.28 |
| bilirubin.glucuronidated+bilirubin.albumin bound | 1968-7 | 0.60 | 1.32 |
| monocytes 2 | 742-7 | 0.75 | 1.34 |
| monocytes 3 | 743-5 | 0.81 | 1.37 |
| eosinophils 2 | 26449-9 | 0.46 | 1.38 |
| alanine aminotransferase | 1742-6 | 0.64 | 1.38 |
| glucose | 2345-7 | 0.66 | 1.38 |
| monocytes/100 leukocytes | 26485-3 | 0.76 | 1.41 |
| sodium 2 | 2947-0 | 0.67 | 1.57 |
| coagulation surface induced | 14979-9 | 0.80 | 1.57 |
| monocytes/100 leukocytes 3 | 744-3 | 0.71 | 1.64 |
| eosinophils/100 leukocytes 2 | 714-6 | 0.65 | 1.65 |
| hematocrit | 4544-3 | 0.50 | 1.76 |
| granulocytes/100 leukocytes 2 | 19023-1 | 0.72 | 1.83 |
| carbon dioxide | 2028-9 | 0.61 | 1.87 |
| glomerular filtration rate/1.73 sq m.predicted | 98979-8 | 0.57 | 1.88 |
| bilirubin | 1975-2 | 0.57 | 1.99 |
| protein 2 | 2885-2 | 0.56 | 2.00 |
| albumin | 1751-7 | 0.53 | 2.02 |
| potassium | 2823-3 | 0.70 | 2.24 |
| sodium | 2951-2 | 0.65 | 2.47 |
| creatinine 2 | 38483-4 | 0.40 | 2.52 |
| granulocytes/100 leukocytes | 30395-8 | 0.71 | 2.59 |
| aspartate aminotransferase | 1920-8 | 0.60 | 2.6 |
| chloride | 2075-0 | 0.63 | 2.69 |
| eosinophils/100 leukocytes | 26450-7 | 0.57 | 2.82 |
| creatinine | 2160-0 | 0.37 | 2.84 |
| magnesium | 19123-9 | 0.60 | 2.96 |
| neutrophils/100 leukocytes | 26511-6 | 0.71 | 3.04 |
| basophils | 704-7 | 0.60 | 3.06 |
| neutrophils/100 leukocytes 2 | 770-8 | 0.73 | 3.09 |
| eosinophils | 712-0 | 0.54 | 3.12 |
| basophils/100 leukocytes 3 | 706-2 | 0.69 | 3.16 |
| basophils/100 leukocytes | 707-0 | 0.73 | 3.21 |
| basophils 2 | 26444-0 | 0.64 | 3.34 |
| basophils/100 leukocytes 2 | 30180-4 | 0.70 | 3.35 |
| gamma glutamyl transferase | 2324-2 | 0.37 | 3.57 |

